# Moving Beyond the ‘Pro’ or ‘Anti’ Binary: Insights into Vaccination Attitudes and Behaviour, with a Case Study from a London, UK Survey

**DOI:** 10.64898/2026.06.29.26356841

**Authors:** Ioannis Rotous, Ysabella-Rozetta Hawkings, Nathan Green

## Abstract

**Background:** Responses to vaccination programmes vary widely, with contrasting perceptions of benefits and harms. This study aimed to provide a contemporary characterization of distinct groups of respondents by moving past traditional ‘pro’ versus ‘anti’ vaccination binaries. The results captured the complex spectrum of vaccine sentiment to directly inform the next strategic phases of a multi-year public health campaign regarding vaccine perceptions and behaviours within the diverse population of London.

**Methods:** Data came from the 2024 *Why We Get Vaccinated* survey conducted in London, UK using a community-based participatory approach. Participants answered vaccine-related questions based on previous vaccine uptake, future willingness to vaccinate (WTV), perceived effectiveness, and safety concerns. We employed Bayesian Binomial logistic regression to identify sociodemographic predictors of individual responses. Subsequently, a Bayesian Nonparametric Latent Class Analysis (BNP-LCA) using a Dirichlet Process Mixture model was used to identify distinct groups and their sociodemographic compositions.

**Results:** The LCA identified five distinct classes. Class 1, the Consistent Uptakers (37.2%), exhibited high vaccine uptake (89.2%) and near-universal belief in efficacy (98.0%) with minimal safety concerns. Class 2, the Concerned Uptakers (17.5%), maintained high WTV (72.2%) despite significant concerns regarding adverse effects (71.2%). Class 3, the Consistent Refusers (18.4%), demonstrated uniform rejection of vaccines and high levels of safety concern (74.4%). Class 4, the Unconcerned Refusers (20.8%), acknowledged vaccine effectiveness (98.4%) but showed very low uptake (1.7%), possibly due to low perceived personal risk rather than active resistance. Class 5, the Undecided/Uninformed (6.0%), was characterized by pervasive uncertainty and “Don’t Know” responses regarding efficacy (80.3%) and vaccination intent. Sociodemographic analysis revealed that residential instability, i.e renting, and caregiving responsibilities were significant barriers to uptake, while older age and retirement were the strongest positive predictors.

**Conclusions:** Vaccine behaviour and attitudes in London are a multidimensional construct. The discovery of “Unconcerned Refusers” and “Concerned Uptakers” highlights that “one-size-fits-all” public health campaigns may not be effective. Tailored public health messaging is required to address the specific uncertainties and socio-economic barriers identified across these distinct profiles. These findings provide a functional framework that will directly guide upcoming resource allocation, messaging adaptations, and policy decisions for the next iterations of the ongoing campaign.

## 1. Introduction

Vaccines have long been established to benefit public health (e.g., controlling infectious diseases such as measles, mumps, rubella (MMR vaccine), influenza, and more recently COVID-19). Despite this, it is common for vaccination coverage to not meet its targets. Some countries have stricter policies but the UK does not enforce vaccinations, rather allowing individuals to opt-in to vaccinations they are eligible for.

The UK Health and Security Agency’s (UKHSA) public perception tracker (PPT) and annual attitudinal survey show that overall, perceptions of vaccinations are positive across the UK^1^. However, in the UK, vaccination coverage decreased in 2023–24 for all of the 14 vaccine coverage measures reported by the NHS Childhood Vaccination Coverage Statistics report. None of these met the 95% target^2^. In particular, London has some of the lowest vaccination rates in the UK. Recent data show a continuing decline with no childhood vaccines, such as MMR, meeting the 95% target set by the WHO to prevent the spread of disease across the population. For example, just 72% of children in London have had two doses of the MMR vaccine by age 5, the lowest of any region and significantly below the national average for England at 84%. As stated in the London Councils vaccination campaign ^3^, “Over a third of London boroughs are even further below with uptake rates below 70%”. Greater London uptake performs especially poorly for influenza vaccination, with coverage 6.6% lower than the national average in 2019–20, a gap wider than it has been since the programme began in 2000.

An individual’s circumstances, feelings and attitudes to vaccines determine the likelihood they will get themselves or their children vaccinated. These can be shaped due to perceptions of ineffectiveness, concerns about harms from side effects and adverse effects, social norms, lack of trust in institutions and health systems, and concerns regarding the underlying reasons for promoting vaccination to the population^4^. The reasons for having these attitudes may include a lack of information, their own previous experiences or those of their friends, family, and community, personality type, and different group membership, and may change over time as their lives and personal circumstances change (e.g., developing a long-term condition or having children).

These varied influences can be systematically understood through the COM-B model of behaviour, which posits that any behaviour requires three interacting components: Capability (psychological and physical), Opportunity, and Motivation^5,6^. To effectively design interventions, we must first understand which of these components are acting as barriers for different segments of the population.

Specifically, in the UK, studies after the COVID-19 pandemic and Britain’s exit from the European Union (“Brexit”) have shown that public trust in institutions has dropped^7–10^. The impact of misinformation and false information, such as the Wakefield MMR publication or negative social media activity, has been shown to reduce uptake of vaccinations, the impact of which can be seen most acutely in the significant increase in measles cases and outbreaks throughout the country in 2024 and 2025^11,12^. This pattern has increased dramatically since the COVID-19 pandemic and has been defined by the World Health Organisation (WHO) ^13,14^ as a global health threat. Since the pandemic, there appears to also be an increase in scepticism about repeated vaccinations, with people questioning the effectiveness of the vaccinations due to the need to be vaccinated annually as with flu and COVID-19^15^.

Due to this current situation, communicating with and informing members of the public in ways that foster trust and positive perceptions is key to improving vaccine uptake throughout the life course. Continued trust in institutions and vaccinations will prevent the re-emergence of infectious diseases and effective management of future outbreaks through higher and more consistent uptake of vaccinations.

It has also been suggested that the success of previous campaigns has reduced the perceived need for vaccinations because the diseases they protect against are now rarely seen^16^. The sense that these diseases are no longer a risk means that people do not recognise the importance of vaccination in keeping this risk low. The work done with regards to the recent increases in measles cases in the UK has included reports of people saying phrases that reflect this sense that measles isn’t a concern any more, and a lack of understanding of how severe measles can be.

To understand and intervene to change attitudes, it is necessary first to identify which groups of individuals have these different views on vaccines. That is, to quantify the associations between sociodemographic characteristics and vaccination related outcomes. It is critical to move beyond binary definitions of simply ‘pro’ or ‘anti’ vaccination and identify the distinct subgroups holding these varying views. Treating public attitude as a simple binary is an oversimplification which can result in misallocated public health resources, alienated demographics, and ineffective outreach. Capturing a multi-dimensional, non-binary reality is therefore an operational necessity for modern public health strategy. While many studies investigate single outcomes separately^17–20^, fewer use combined approaches, such as *Latent Class Analysis (LCA)*, to identify distinct groups of vaccine attitudes across multiple dimensions. LCA can help address the previous gap by identifying groups with shared beliefs and behaviour patterns which can inform approaches to communicating and engaging groups with lower vaccination uptake.

An important attitude is often called vaccine *hesitancy* and identifying a vaccine hesitant group could be of particular benefit. This is an indecision or reluctance toward vaccination. Choi Y *et al*. (2025) ^21^ investigated vaccine-hesitant subgroups in 12 Western Pacific countries in 2021-22 using LCA. Their analysis identified nine latent classes, which they distinguished by combinations of formal education, age and employment status. They highlighted that the probabilities of COVID-19 vaccine acceptance and booster uptake were significantly lower in most of these latent classes, compared to university-educated older adults. There are several examples of LCA applied to COVID-19 vaccine attitudes^22,23^ in the US. For vaccine hesitancy and mistrust, in particular,^24^ found five latent classes they called “High Trust”, “Trust in Own Doctor”, “High Distrust”, “Undecided” and “No Opinion”. They found a non-linear association between age groups and willingness to vaccinate (WTV). Relative to those aged 65 and older, those in the 35–54 age group were less likely to be vaccinated. Women appeared 10% less likely than men to plan to get vaccinated. Willis et al. (2025)^25^ investigated vaccine hesitancy for vaccines generally, COVID-19 and also HPV. They used online survey data from parents of paediatric patients recruited through eight clinics within the University of Arkansas for Medical Sciences Rural Research Network in 2022. They found three latent classes they called “Selectively Hesitant”, “COVID-Centric Hesitant,” and “Pervasively Hesitant.” Significant predictors of class membership included age and education. In the UK, there appear fewer such examples. Using original survey data collected in the US, UK, and Canada, Gravelle et al. (2022) ^26^ found five latent classes from strong supporter to “anti-vaxer” with a fifth “error” class. They highlight education and political leaning as notably different between classes.

The primary aim of this work is to identify distinct classes of vaccine attitudes and behaviours using data from a 2024 London-wide survey. Secondary aims include describing the characteristics (response patterns to vaccine questions) of these classes, and to examine the association between sociodemographic factors and membership in the identified classes. Understanding these distinct groups can inform the development of more targeted and effective public health communication strategies and interventions to improve vaccine confidence and uptake.

## 2. Methods

### 2.1. Study Design and Participants

The pan-London *Why We Get Vaccinated* (WWGV) campaign is a three-year campaign created with local communities, UKHSA, the NHS, London Borough Councils and the London regional Public Health communications network of the Association of Directors of Public Health (ADPH) London; the organisation acts as a coordinating body, championing public health across all of London’s 32 boroughs and the City of London, and providing a forum for the Directors of Public Health (DsPH) to collaborate.

In 2024, YouGov conducted an online survey, as part of the WWGV campaign, among adults aged *≥*18 living across the 32 London boroughs and the City of London. Fieldwork was undertaken between 6th–12th August 2024. The survey included both closed (multiple choice) and open-ended questions and was conducted using YouGov’s online panel of individuals who have voluntarily agreed to participate in research. A quota-based, non-probability sampling approach was used to ensure the sample reflected the demographics of the campaign’s target population. Data were subsequently weighted to be representative of all London adults (aged *≥*18), based on age, gender, socio-economic status, and geographic distribution. Ethical approval was obtained for the original survey, and informed consent was secured from all participants.

### 2.2. Survey Questions and Covariates

We analysed participants’ responses to three questions and two statements designed to elicit their perspectives on various aspects of vaccination. In latent class terminology, these are called *manifest* variables. These were

Q1. Did you get the flu vaccine last winter? (Yes, I did; No, I didn’t; Don’t know/can’t recall)

Q2. How likely, if at all, are you to get the flu vaccine this year? (Not very likely; Fairly likely; Don’t know/unsure; Not at all likely; Very likely)

Q3. Have you vaccinated your oldest child with one or more doses of the MMR vaccine? (No, I haven’t; Don’t know/unsure; Yes, I have; Not applicable - My oldest child is still not old enough to get the MMR vaccine)

Q4. Vaccines are effective (Strongly disagree; Tend to disagree; Neither agree nor disagree; Tend to agree; Strongly agree)

Q5. I am concerned about serious adverse effects (i.e., negative side effects or unwanted results) of vaccines (Strongly disagree; Tend to disagree; Neither agree nor disagree; Tend to agree; Strongly agree)

The survey collected several sociodemographic characteristics known to influence vaccination attitude and behaviours^27,28^, including: Ethnicity (White, Asian, Black, Mixed, Other, Prefer not to say), Age group years (18–24, 25–34, 35–44, 45–54, *≥* 55), Gender (Male, Female), Social grade (ABC1, C2DE), Marital status (Married/Civil Partnership, Living as married, Never married, Widowed, Separated/Divorced, Missing), Working status (Working full-time, Working part-time, Full-time student, Retired, Unemployed, Not working/Other), Number of Children in household (0, 1, 2, *≥* 3, Refused),

Parent/Guardian age group of dependent years (*≤* 4, 5–11, 12–16, 17–18, *>* 18 years, Not parent/Guardian), House tenure (Rent from a local authority, Own (with a mortgage or shared ownership scheme or outright), Rent from a private landlord, Live rent-free with my parents, Live with parents but pay some rent to them, family or friends, Rent from a housing association, Other, Missing), Gross household and Gross personal income bands, Social media messaging service use in the last month (Yes, No from eight choices). For our analysis, we did not use the income or social media variables. Income is strongly correlated with the other variables included, and the social media service is weakly informative for the number of variables. In the future, a derived single variable could be considered.

The social grade terms of ABC1 (High) and C2DE (Low) refer to social classifications used primarily in the UK for market research and audience segmentation. They are based on occupation and were originally developed by the National Readership Survey (NRS) ^29^. Specifically, ABC1 is often used to refer to more affluent or middle-class groups (Grades A, B, and C1), and C2DE is often used to refer to less affluent or working-class groups (Grades C2, D, and E).

Missing data were explicitly coded as missed, whereas individuals who refused to answer were coded as refused.

### 2.3. Statistical Analysis

Descriptive statistics were calculated for all variables and are shown in Table 1.

**Table 1:**
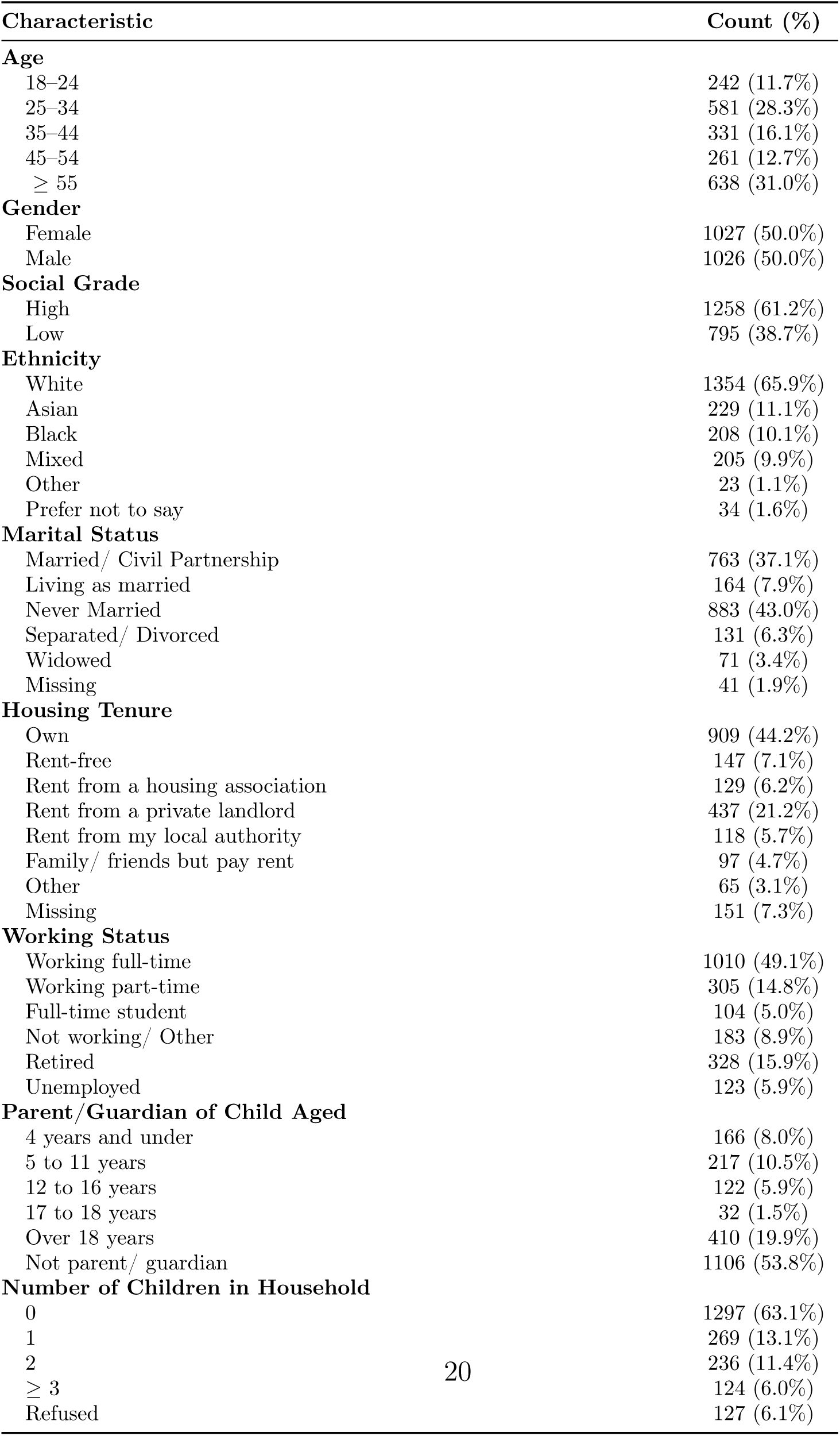
Demographic total counts and percentage statistics from the 2024 London “Why We Get Vaccinated” survey.

| Characteristic | Count (%) |
| --- | --- |
| **Age** |  |
| 18–24 | 242 (11.7%) |
| 25–34 | 581 (28.3%) |
| 35–44 | 331 (16.1%) |
| 45–54 | 261 (12.7%) |
| ≥ 55 | 638 (31.0%) |
| **Gender** |  |
| Female | 1027 (50.0%) |
| Male | 1026 (50.0%) |
| **Social Grade** |  |
| High | 1258 (61.2%) |
| Low | 795 (38.7%) |
| **Ethnicity** |  |
| White | 1354 (65.9%) |
| Asian | 229 (11.1%) |
| Black | 208 (10.1%) |
| Mixed | 205 (9.9%) |
| Other | 23 (1.1%) |
| Prefer not to say | 34 (1.6%) |
| **Marital Status** |  |
| Married/ Civil Partnership | 763 (37.1%) |
| Living as married | 164 (7.9%) |
| Never Married | 883 (43.0%) |
| Separated/ Divorced | 131 (6.3%) |
| Widowed | 71 (3.4%) |
| Missing | 41 (1.9%) |
| **Housing Tenure** |  |
| Own | 909 (44.2%) |
| Rent-free | 147 (7.1%) |
| Rent from a housing association | 129 (6.2%) |
| Rent from a private landlord | 437 (21.2%) |
| Rent from my local authority | 118 (5.7%) |
| Family/ friends but pay rent | 97 (4.7%) |
| Other | 65 (3.1%) |
| Missing | 151 (7.3%) |
| **Working Status** |  |
| Working full-time | 1010 (49.1%) |
| Working part-time | 305 (14.8%) |
| Full-time student | 104 (5.0%) |
| Not working/ Other | 183 (8.9%) |
| Retired | 328 (15.9%) |
| Unemployed | 123 (5.9%) |
| **Parent/Guardian of Child Aged** |  |
| 4 years and under | 166 (8.0%) |
| 5 to 11 years | 217 (10.5%) |
| 12 to 16 years | 122 (5.9%) |
| 17 to 18 years | 32 (1.5%) |
| Over 18 years | 410 (19.9%) |
| Not parent/ guardian | 1106 (53.8%) |
| **Number of Children in Household** |  |
| 0 | 1297 (63.1%) |
| 1 | 269 (13.1%) |
| 2 | 236 (11.4%) |
| ≥ 3 | 124 (6.0%) |
| Refused | 127 (6.1%) |

The full analysis was performed in two steps. First, to understand the relationship between sociodemographic factors and individual vaccine attitudes and behaviour, we conducted a series of Bayesian Binomial logistic regression analyses fitted separately for each of the five key outcome questions and statements. Responses were grouped into either positive or negative. Individuals who gave responses that were neither positive nor negative were dropped from this analysis. For Q1, Q2, and Q3, responses were already in this form. For Q4 and Q5, which were on a Likert scale, responses were merged into positive groups {Strongly agree, Tend to agree}, {Very likely, Fairly likely} and negative groups {Strongly disagree, Tend to disagree}, {Not very likely, Not at all likely}, respectively. The results of this analysis will indicate variables of interest for each question, which will complement and validate the LCA which considers all questions simultaneously.

Following the preliminary analysis of categorical effects on items Q1–Q5, we employed a Bayesian Nonparametric Latent Class Analysis (BNP-LCA), with the use of a Dirichlet Process Mixture model^30,31^, to identify groups of individuals who respond similarly to Q1–Q5 and can be classified into classes reflecting similar vaccination attitudes and behaviours. In most LCA studies, the number of groups is subjectively pre-specified. It is not inferred from the data but determined a priori by the researcher. In contrast, the BNP-LCA framework treats the number of groups as a random parameter that is inferred from the joint responses to Q1–Q5. This BNP-LCA approach yields a more granular and data-driven representation of the underlying population structure, able to explore classes that often are missed^32,33^.

To enhance model identifiability and minimize the risk of over-fitting, we collapsed rare response categories for items Q1–Q5 prior to analysis. This prevents the model from generating meaningless or redundant classes simply because certain combinations of answers are extremely rare in the data. By consolidating these low-frequency observations, we ensure that the resulting profiles reflect genuine, generalizable patterns of behaviour rather than random sampling noise. Specifically, we collapsed the response scales for Q2, Q4, and Q5. For Q2, responses were pooled into three ordinal categories of {Not likely, Don’t know, and Likely}. Similarly, for Q4 and Q5 responses were pooled into {Disagree, Neither agree nor disagree, and Agree}.

We characterized the response profile of each latent class using the posterior mode. In practical terms, this means each class is summarized by identifying the most probable answer its members would give to every question and statement in the model. After the identification of the classes, a Bayesian Multinomial logistic regression was conducted to create distinct individual profiles for each latent class. This stage of the analysis sought to associate class-specific vaccination behaviours and attitudes with the socioeconomic and demographic characteristics of individuals. By doing so, we aimed to identify the key determinants that influence vaccination attitudes observed across the latent profiles. Uncertainty about class membership was propagated from the LCA using the BCH approach^34^.

Appropriate LCA analysis and reporting guidelines were followed^35^. The code is available at https://github.com/n8thangreen/vaccineanalysis.

## 3. Results

### 3.1. Study Data

The sociodemographic characteristics of the sample are presented in Table 1. In total, there were *N* = 2053 survey responses. The median age group was 45–54 years, and 50.0% identified as female, 61.2% belonged to the high social grade. The majority of individuals were never married (43.0%). The remainder were married or in a civil partnership (37.1%), living as married (7.9%), separated/divorced (6.3%), and widowed (3.4%). For housing, the highest proportion of people owned their home outright (44.2%). The next most frequent group rents from a private landlord (21.2%) or from a local authority (5.7%). Approximately half the respondents worked full-time (49.1%), with the next largest group being retired (15.9%) or working part-time (14.8%). The most common parent/guardian situation was to not have children (53.8%) followed by have adult children (19.9%), 5 to 11 year old (10.5%), 4 years and under (8.0%), and 12 to 16 years (5.9,%). For the number of children in the household, 63.1% responded none, implying that adult children do not live at home, consistent with the 31.0% aged 55 and over.

Complete response distributions for all vaccine attitude questions are given in the Appendix.

### 3.2. Separate Regression Analyses

#### 3.2.1. Q1: Did you get the flu vaccine last winter?

The relative strength of the associations in terms of the posterior ORs is shown in Figure 1. Several sociodemographic factors demonstrated a substantial negative impact on vaccine uptake last winter. Notably, homeowners exhibited significantly higher odds of vaccination compared to non-homeowners. These findings were compounded by negative associations in age groups 25–34 and 45–54 and among parents or guardians for children of all ages. This might suggest that residential precariousness and caregiving demands may represent significant structural barriers to vaccination access or intent.

**Figure 1:**
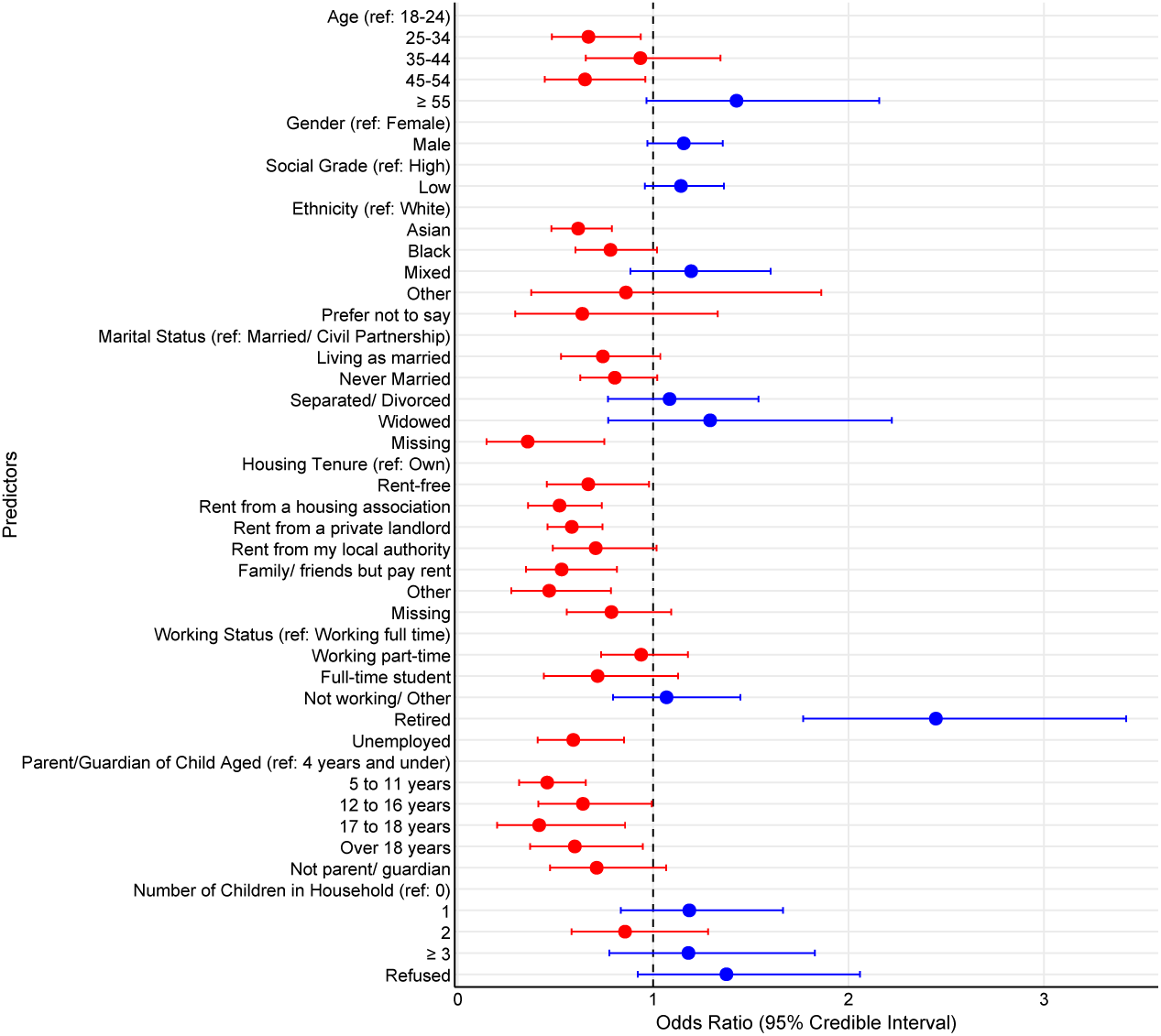
Forest plot of Odds Ratios (95% credible intervals) for the WWGV survey question responses to did you get the flu vaccine last winter. Blue and red colours indicate an increase in the odds (OR *>* 1) and decrease in the odds (OR *<* 1), respectively.

In contrast, the results suggested that socio-economic stability and life stage were primary drivers of uptake. Retirement and older age (*≥* 55) exhibited the strongest positive associations, with the “Retired” status showing the biggest increase in odds. Furthermore, increased odds of uptake were observed among males, individuals of low socio-economic status, and those within the mixed ethnic group.

#### 3.2.2. Q2: How likely, if at all, are you to get the flu vaccine this year?

The posterior ORs are shown in Figure 2. Several factors were associated with significantly reduced odds of WTV. In particular, individuals in the 45–54 age cohort and those renting from a housing association or a private landlord exhibited a markedly lower likelihood of intending to vaccinate. Furthermore, parental or guardian responsibilities were consistently aligned with reduced odds.

**Figure 2:**
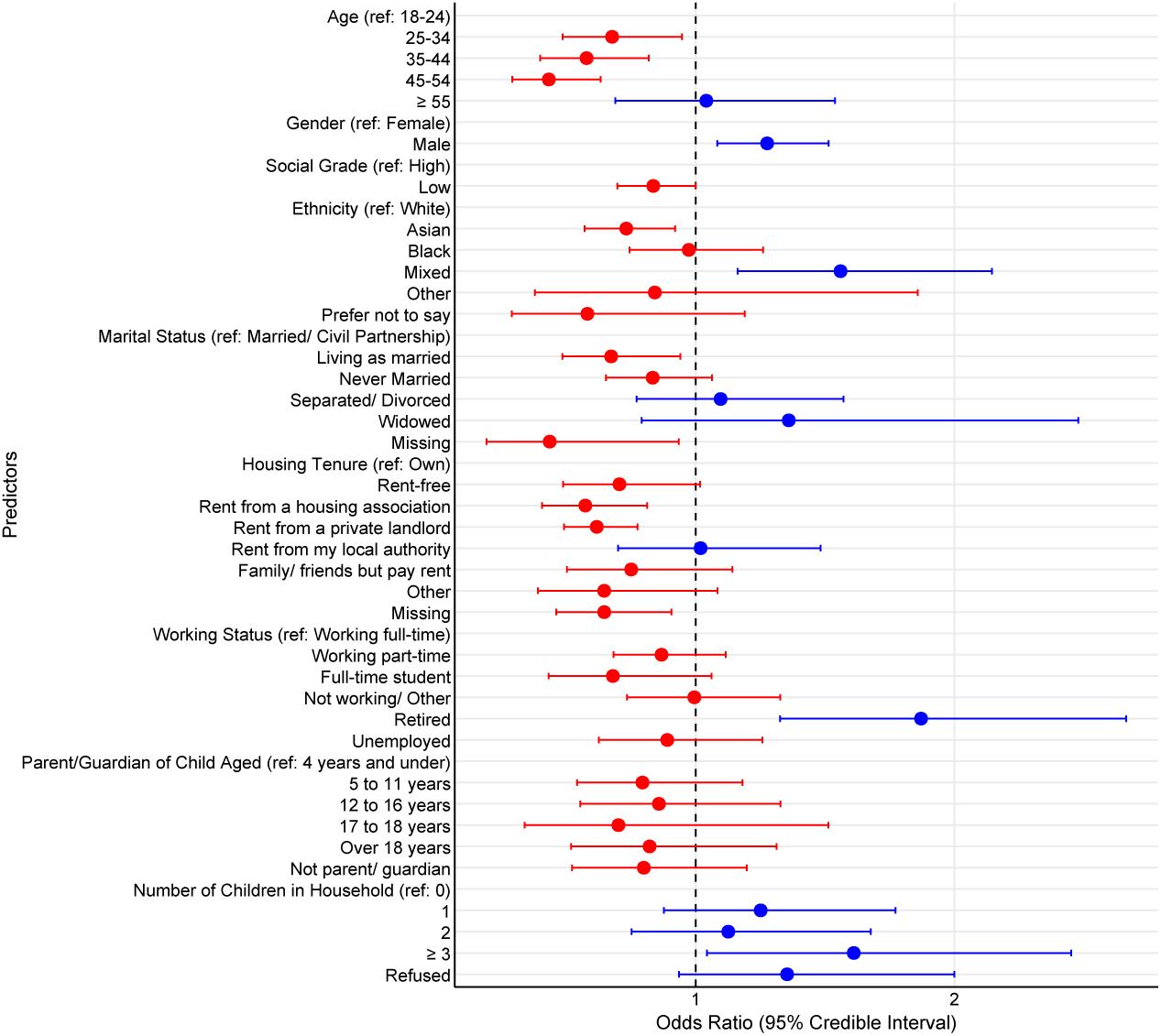
Forest plot of Odds Ratios for flu vaccination intent next year (95% credible intervals). Blue and red colours indicate an increase in the odds (OR *>* 1) and decrease in the odds (OR *<* 1), respectively.

The higher odds of WTV were positively associated with being male, identifying as mixed ethnicity, being widowed, or being retired. Notably, the presence of multiple children in the household also appeared to correlate with increased intent.

#### 3.2.3. Q3: Have you vaccinated your oldest child with one or more doses of the MMR vaccine?

The posterior ORs are visualized in Figure 3. The most substantial negative predictor was full-time student status, which was associated with significantly reduced odds of MMR uptake versus working full-time individuals. Also, living rent-free was associated with reduced odds of MMR uptake compared to individuals who own their household. Other predictors that showed small odds included various rental and housing tenures, such as private rentals and housing associations. Additionally, several parental or guardian responsibilities and younger age cohorts (notably the 25–34 group) showed a similar downward trend.

**Figure 3:**
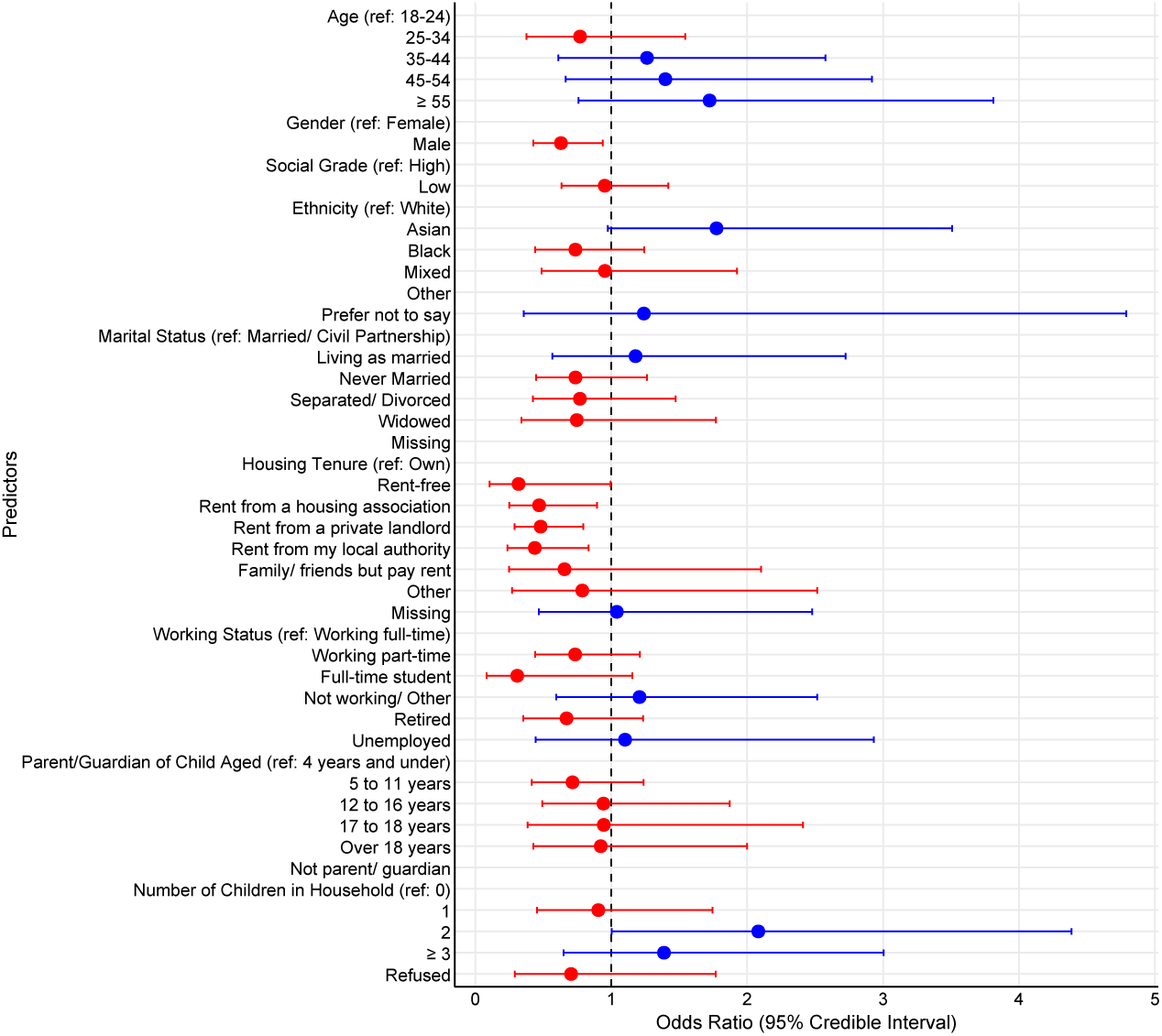
Forest plot of Odds Ratios for child’s MMR vaccination history (95% credible intervals). Blue and red colours indicate an increase in the odds (OR *>* 1) and decrease in the odds (OR *<* 1), respectively.

Increased odds of MMR vaccination uptake were notably associated with individuals aged 55 and over, and those of Asian ethnicity. Furthermore, higher WTV was observed among those who own homes or those who have more than two children.

#### 3.2.4. Q4: Vaccines are effective

The posterior ORs are shown in Figure 4. Most of the factors were positively associated with the belief that vaccines are effective. Nevertheless, several key factors were associated with significantly reduced odds of perceived efficacy. The residential factors of renting from a local authority or housing association had the largest negative associations. Marital status was also a significant predictor; individuals who were separated, divorced, or widowed showed a markedly lower likelihood of perceiving vaccines as effective compared to other groups. Furthermore, decreased perceptions of efficacy were observed among the 35–44 and 45–54 age groups, as well as individuals in lower social grades.

**Figure 4:**
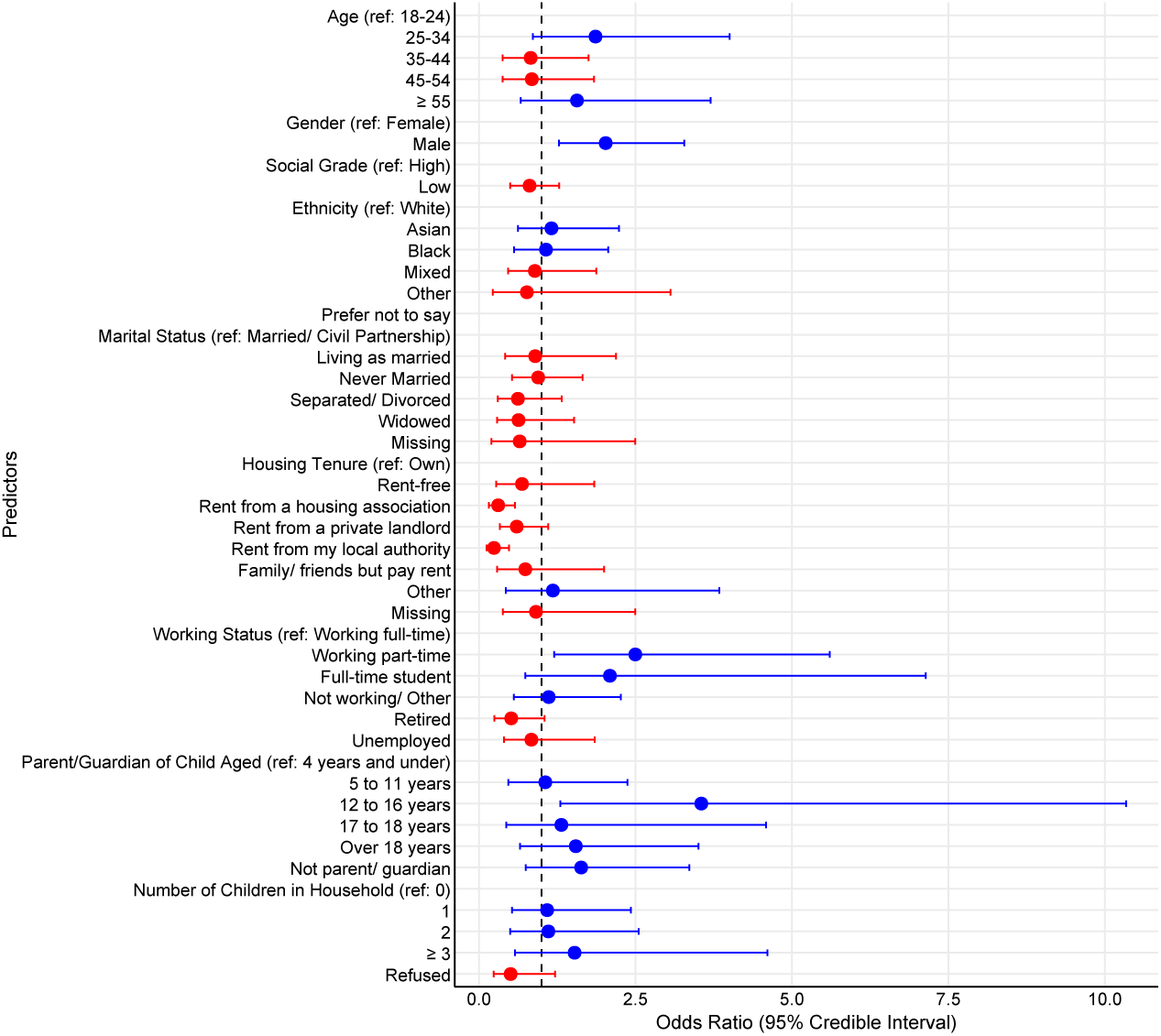
Forest plot of Odds Ratios for perceived vaccine effectiveness (95% credible intervals). Blue and red colours indicate an increase in the odds (OR *>* 1) and decrease in the odds (OR *<* 1), respectively.

#### 3.2.5. Q5: I am concerned about serious adverse effects of vaccines

The posterior ORs are shown in Figure 5. Several factors were associated with a significant reduction in the odds of concern, most notably the age *≥* 55 cohort, followed by the 35–44 and 45–54 years. Other predictors that showed diminished odds of concern included individuals who do not have parental or guardian responsibilities at the present time. These results demonstrate that, while safety concerns are statistically concentrated within specific household and ethnic groups, older age cohorts and non-parental groups consistently displayed a lower likelihood of concern regarding adverse vaccine effects.

**Figure 5:**
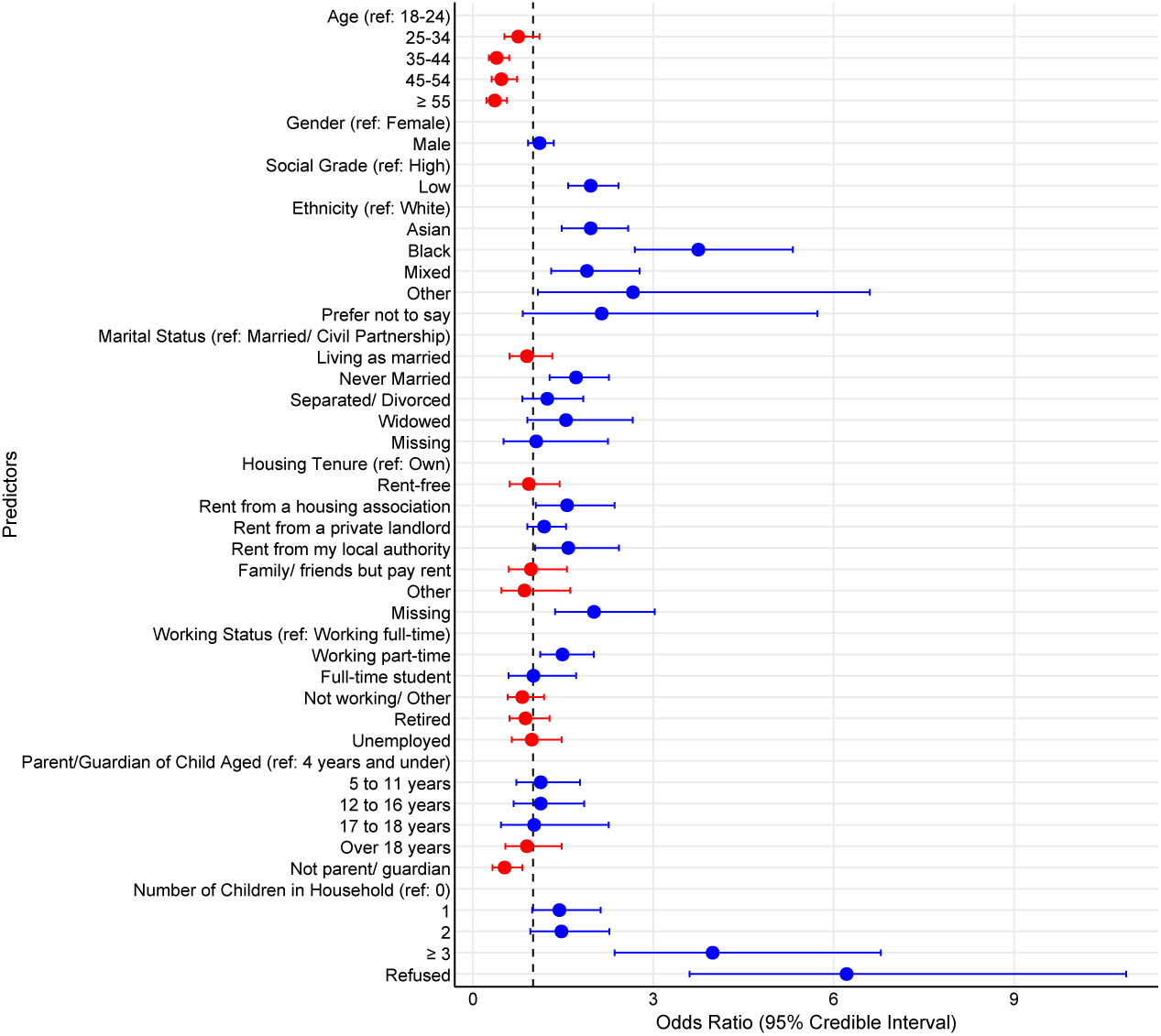
Forest plot of odds ratios for concern about serious adverse side effects (95% credible intervals). Blue and red colours indicate an increase in the odds (OR *>* 1) and decrease in the odds (OR *<* 1), respectively.

Comprehensive posterior summaries of covariate log ORs coefficients and credible intervals for all regression models are detailed in the Appendix.

### 3.3. Model Validation

Finally, the goodness-of-fit for each Bayesian logistic regression model was evaluated using Posterior Predictive Checks (PPC)^36^. As illustrated in the Appendix, the observed data were consistently recovered by the posterior predictive distributions. The absence of systematic discrepancies across these checks confirmed that the models were adequately specified and effectively captured the underlying data distributions.

### 3.4. Latent Class Analysis

The estimation of the BNP-LCA inferred that a solution with 5 classes best represents respondent attitude and behaviour across the vaccination survey. These classes characterize the response patterns for Q1, Q2, Q4, and Q5 (excluding Q3 regarding MMR child vaccination). The general pattern of probabilities within and between the classes is summarised in Table 2.

**Table 2:** Within-class posterior class membership probabilities (mean and standard deviation) estimated using a Latent Class Analysis model and data from the “Why We Get Vaccinated” Survey (2024). Bold values indicate the posterior mode estimate for the most probable answer per question.

|  |  |  | Latent Classes |  |  |  |  |
| --- | --- | --- | --- | --- | --- | --- | --- |
| Question |  | Response Category | Class 1<br>Consistent Uptakers | Class 2<br>Concerned Uptakers | Class 3<br>Consistent Refusers | Class 4<br>Unconcerned Refusers | Class 5<br>Undecided / Uninformed |
| 1 | Did you get the flu vaccine last winter? | Don't know | 0.005 (0.004) | 0.241 (0.093) | 0.007 (0.006) | 0.036 (0.012) | 0.400 (0.111) |
|  |  | No, I didn't | 0.102 (0.044) | 0.345 (0.133) | **0.943** (0.021) | **0.947** (0.018) | **0.411** (0.139) |
|  |  | Yes, I did | **0.892** (0.045) | **0.414** (0.104) | 0.050 (0.020) | 0.017 (0.014) | 0.189 (0.080) |
| 2 | How likely, if at all, are you to get the flu vaccine this year? | Don't know | 0.006 (0.004) | 0.227 (0.083) | 0.026 (0.017) | 0.185 (0.028) | **0.521** (0.128) |
|  |  | Unlikely | 0.018 (0.009) | 0.051 (0.042) | **0.918** (0.051) | **0.705** (0.057) | 0.413 (0.141) |
|  |  | Likely | **0.977** (0.010) | **0.722** (0.089) | 0.056 (0.045) | 0.110 (0.067) | 0.066 (0.059) |
| 3 | Have you vaccinated your oldest child with the MMR vaccine? | Don't know | 0.028 (0.006) | 0.068 (0.026) | 0.051 (0.014) | 0.007 (0.005) | 0.108 (0.047) |
|  |  | No, I haven't | 0.029 (0.007) | 0.096 (0.038) | 0.106 (0.020) | 0.016 (0.008) | 0.023 (0.022) |
|  |  | Not applicable | **0.497** (0.019) | **0.587** (0.057) | **0.579** (0.032) | **0.627** (0.029) | **0.798** (0.067) |
|  |  | Yes, I have | 0.446 (0.020) | 0.248 (0.055) | 0.264 (0.032) | 0.350 (0.028) | 0.071 (0.051) |
| 4 | Vaccines are effective | Neither | 0.015 (0.008) | 0.167 (0.054) | 0.310 (0.048) | 0.010 (0.009) | **0.803** (0.123) |
|  |  | Tend to disagree | 0.005 (0.003) | 0.025 (0.014) | 0.120 (0.023) | 0.005 (0.005) | 0.025 (0.024) |
|  |  | Tend to agree | **0.980** (0.008) | **0.809** (0.057) | **0.570** (0.060) | **0.984** (0.010) | 0.171 (0.122) |
| 5 | Concerned about negative side effects | Neither | 0.209 (0.016) | 0.239 (0.068) | 0.224 (0.051) | 0.240 (0.037) | **0.823** (0.099) |
|  |  | Tend to disagree | **0.491** (0.026) | 0.049 (0.034) | 0.032 (0.017) | **0.616** (0.069) | 0.028 (0.028) |
|  |  | Tend to agree | 0.301 (0.025) | **0.712** (0.075) | **0.744** (0.052) | 0.144 (0.069) | 0.149 (0.094) |
| Total |  |  | 0.372 (0.203) | 0.175 (0.250) | 0.184 (0.173) | 0.208 (0.175) | 0.060 (0.110) |

In the following, we introduce the labels assigned to each class and define their characteristics. These labels reflected the predominant behavioural patterns of the members within each respective group.

Class 1 (Consistent Uptakers) constituted the most pro-vaccination group, characterized by high uptake and lack of safety concerns. Class 2 (Concerned Uptakers) represented individuals who, despite a history of vaccination and future WTV, maintained significant concerns regarding negative effects, suggesting a mindset of compliance but with caution. Class 3 (Consistent Refusers) showed a uniform rejection of vaccination, reporting no past or future WTV along with scepticism about safety. Class 4 (Unconcerned Refusers) acknowledged vaccine efficacy but lacked a WTV, indicating a gap where positive beliefs do not translate into health actions. Class 5 (Undecided/Uninformed) was defined by pervasive uncertainty, with respondents consistently opting for “Don’t Know” or “Neutral” responses, indicating a lack of engagement with vaccination discourse rather than stated opposition. The five LCA behavioural and attitudinal profiles illustrate a complex landscape of vaccine engagement. The largest group, Consistent Uptakers (37.2%), served as the baseline for high adherence, characterized by nearly universal vaccine uptake (89.2%) and high perceived efficacy (98.0%). Conversely, Consistent Refusers (18.4%) demonstrated a pattern of vaccine avoidance; despite a moderate acknowledgment of vaccine effectiveness (57.0%), their decisions appeared to be dominated by the highest levels of concern (74.4%) and a nearly complete absence of past or future WTV.

A notable finding was the emergence of Unconcerned Refusers (20.8%), who did not appear to take up vaccination despite their beliefs. Although this group aligns with the Uptakers group in terms of vaccine effectiveness (98.4%), they exhibited very low vaccination rates (1.7%) and future TVW (11.0%). Given their low levels of concern about negative side effects(14.4%), their lack of uptake may be due to a lack of perceived personal risk rather than active resistance. In contrast, Concerned Uptakers (17.5%) appeared highly engaged but conflicted; they report high levels of concern (71.2%) but showed vaccination intent (72.2%), significantly higher than their behaviour in their previous year (41.4%).

Finally, the Undecided/Uninformed class (6.0%) represented a small but distinct segment of disengagement. Defined by high frequencies of responses “Neither” and “Don’t know”, particularly regarding efficacy (80.3%) and future WTV (52.1%).

#### 3.4.1. Class Composition by Covariates

To investigate the drivers of class membership, we examined the associations between individual covariates and the inferred latent structures. Following the three-step approach proposed by Bolck et al. (2004) ^37^, latent class assignments were regressed on a set of sociodemographic predictors. This framework explicitly accounted for the probabilistic nature of class membership by incorporating a correction for classification error. By doing so, the potential for attenuation bias and inflated standard errors is mitigated, which typically occurs when treating posterior allocations as observed data.

Before fitting the multinomial regression, we collapsed the marital status categories of “Separated/Divorced” and “Widowed” into a single group. Similarly, the parent/guardian dependencies, from 4 years and under to 18 years, were aggregated into a single “*≤* 18” category. The posterior ORs are shown in Figure 6, divided into demographics, economic, and family factors. The corresponding profiles of individuals belonging to each class can be assessed. Consistent Uptakers class is predominantly defined by an older, socio-economically stable demographic. Members of this class are significantly more likely to be aged 55 years or older, and retired, with a high probability of owning a home and being parents of adult children (*>* 18 years). In contrast, this group is characterized by a notable absence of younger age cohorts (25–44) and a significantly lower likelihood of being unemployed or belonging to a lower social grade. This profile suggested a demographic of established individuals with high historical vaccine compliance and fewer economic or household barriers to access to healthcare.

**Figure 6:**
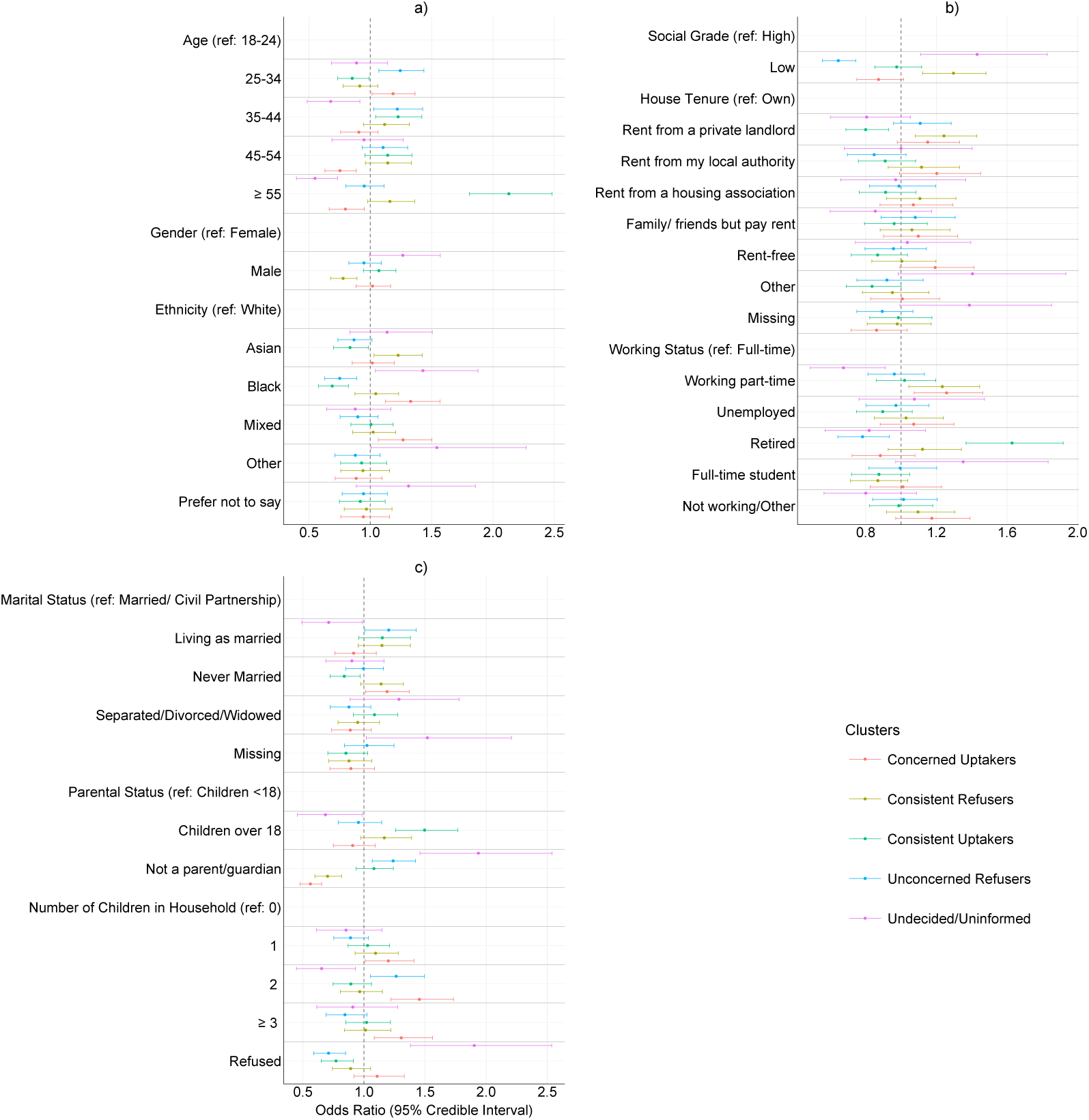
Odds ratios and 95% credible intervals for predictors of latent class membership. The vertical dashed lines indicate where a predictor has no effect, and horizontal lines separate individual covariate levels. Results are grouped by a) demographic predictors (Age, Gender, and Ethnicity), b) socio-economic predictors (Social Grade, House Tenure, and Working Status), and c) family and household composition (Marital Status, Parental Status, and number of children). Reference categories are indicated in the respective headers.

Concerned Uptakers class is uniquely identified by its younger age group (25–44 years) and a strong association with Black and Mixed ethnicities. This association potentially reflects the impact of inequality and lack of institutional trust, often due to negative experiences, that these communities might face. This is the class with the most family members (high odds of having two or more children in the household). Their socio-economic markers are distinct, showing a high reliance on housing associations or private landlords for tenure. Critically, this group is characterized by the near-total absence of retired individuals and those in the *≥* 55 age group. The compliance with caution interpretation of this class likely reflects the complexities of managing health decisions within younger, multi-child households in rented accommodations.

The profile for Consistent Refusers class indicates a demographic that contains more males, and Asian or Other ethnic backgrounds. Professionally, individuals in this class show a greater propensity for part-time employment. A defining negative association for this group is their household structure; they are significantly less likely to be parents or guardians of children under 18 and are under-represented among married or separated/divorced/widowed individuals. Furthermore, they show lower odds of homeownership compared to the baseline. This suggests a more mobile or less family-based demographic whose vaccine rejection may be rooted in individualistic rather than household-centred concerns.

Individuals in Unconcerned Refusers class are predominantly middle-aged, married and likely to own a home with a mortgage, often parents or guardians of children under 18. What distinguishes this group is the significance of lower social grades and unemployed status, suggesting a relatively affluent or professional profile. They are less likely to be from Black or Mixed ethnicity.

Undecided/Uninformed class is defined by socio-economic vulnerability and lack of participation. It shows the strongest association with lower social grades, unemployment, and full-time student status. This class is also marked by a high frequency of “Other” or “Prefer not to say” responses regarding ethnicity and marital status. In particular, this group is defined by what it lacks: it has significantly lower odds of home ownership, parental status, or the *≥* 55 age group.

A significant advantage of our Bayesian non-parametric framework is the flexibility it has for the post-hoc aggregation of latent profiles. This allows for a nested interpretation of the data; while the five-class model provides granular detail, a more parsimonious three-class solution can be derived to distinguish broadly between “pro-vaccination”, “anti-vaccination”, and “un-informed” cohorts. By merging profiles that have shared demographic and attitudinal characteristics, we can create a higher-level strategic perspective for public health intervention.

The Uptakers cohort represents the segment of the population with the highest historical and future commitment to vaccination, although it has many demographic fractions. By merging the “Consistent Uptakers” with the “Concerned Uptakers,” this group spans the age spectrum from young parents (25–44) to retirees (*≥* 55). Socio-economically, this cohort is defined by a high density of multiple child households and a mix of homeowners and social renters, particularly those associated with housing associations. Despite the “caution” noted in the younger members, the unifying characteristic of this cohort is action over apathy; they are the primary participants in vaccination programs. Their defining profile suggests that vaccine uptake is driven by a combination of established social habits (among older individuals) and protective parental responsibility (among younger families).

The difference between intent and action distinguishes the “Consistent Refusers” with the “Unconcerned Refusers”. Collectively, they represent a group where vaccination is either ideologically rejected or practically deprioritized. They are predominantly middle-aged and married, with a strong association with homeownership and part-time employment. A key ethnic signal is the over-representation of Asian and Other groups. The synthesis of these classes reveals a significant psychological and behavioural barrier. While members may acknowledge vaccine efficacy (as in Unconcerned Refusers) or value individual autonomy (as in Consistent Refusers), this does not translate into WTV, and possibly health-seeking behaviour generally. This group is characterized by their lack of uptake, which is due to choice rather than as a result of socio-economic marginalization.

The disengaged cohort has a distinct profile. It is uniquely defined by severe socio-economic precariousness, with the highest chances of unemployment, student status, and belonging to lower social grades. Unlike the other cohorts, which show clear positive or negative intent, this group is defined by neutrality and data gaps, frequently responding with “Don’t Know” or “Refused” regarding ethnicity, marital status, and household composition. They are significantly less likely to own a home or be parents of adult children.

## 4. Discussion

Our study identified five distinct latent classes of vaccine attitudes and behaviours in London, challenging the binary “pro-vaccine” versus “anti-vaccine” paradigm and revealing a complex landscape of engagement. The heterogeneity of these profiles confirms that naive public health campaigns that distinguish individuals in binary classes are likely to be insufficient.

The “Unconcerned Refuser” class, which acknowledged vaccine efficacy (98.4%) but had very low uptake (1.7%), suggests that for a significant portion of the population, the barrier is a lack of perceived personal risk or population-level impact, rather than “anti-vax” sentiment or active resistance. That is, in terms of the COM-B framework, they lack the motivation to act. For this group, because their barrier is not capability, it may be that education is less critical than making vaccination more convenient and investigating “nudging” interventions. In contrast, the “Concerned Uptaker” profile reveals individuals who maintain WTV (72.2%) despite having significant concerns about adverse effects (71.2%). This indicates a need for targeted reassurance and safety messaging to prevent these compliant yet cautious individuals from drifting into refusal. Within the uptaker classes, there may be further distinction which may influence messaging. It has been shown that individuals who felt pressured to vaccinate are positioned between unvaccinated and those who vaccinated voluntarily in their perceptions and intentions^38^. A third group, identified as the “Undecided/Uninformed” class, is defined by significant socioeconomic vulnerability. It is plausible that their engagement with the study was disproportionately driven by extrinsic motivation—specifically, the financial remuneration offered upon survey completion. This introduces the risk of survey satisficing. In this case, responses may be less reflective of genuine attitudes. Within the COM-B framework, this points to a fundamental deficit in psychological capability and opportunity, highlighting the structural and cognitive barriers this demographic faces regarding vaccine literacy.

A consistent theme across the separate Bayesian regression models was the role of socio-economic precariousness as a primary barrier to uptake. Residential instability—specifically renting from private landlords or housing associations—served as a robust negative predictor of both historical uptake and belief in vaccine effectiveness. These findings, coupled with the lower intent observed among individuals with caregiving responsibilities, suggest a so-called “inverse care law” where those in the most deprived circumstances face the highest structural and logistical barriers to accessing preventative health interventions. Extensive research in the UK has consistently demonstrated this clear link between socio-economic hardship and lower vaccination uptake^39^. This aligns with a broader understanding of health disparities, where deprived populations experience poorer health outcomes and lower engagement with preventative services^40^, with London having particularly pronounced disparities related to socio-economic factors^41^. National surveys indicate that this is often linked to lower levels of educational attainment and household income, which correlate with lower health literacy, reduced ability to critically evaluate health information, and diminished trust in scientific expertise .

Conversely, socio-economic stability—characterised by retirement, older age (*≥* 55), and home ownership—remained the strongest association with positive vaccine sentiment and behaviour. However, demographic nuances and local contexts remain critical. We found that Asian and Black ethnicities were associated with different classes (Refusers vs. Concerned Uptakers), highlighting the need to explicitly state that a minority ethnic group is not a homogeneous single group in vaccine research. Furthermore, the specific nature of socio-economic hardship and its impact on health behaviours varies between communities; studies might uncover unique local drivers not captured in larger-scale research, such as the role of specific community leaders, local health service provision, or the impact of highly localised misinformation campaigns. Addressing the systemic exclusion of these marginalized populations is essential to meeting WHO childhood vaccination targets, which currently remain unmet in all measures in London.

### 4.1. Comparison with Previous Work

Our findings corroborate broader evidence investigating single outcomes. A European analysis of 20000 interviews found that upper-class status and tertiary education were significantly associated with positive individualistic and altruistic views on vaccination^17^, mirroring our finding that these groups are more likely to vaccinate and believe in vaccine effectiveness. Similarly, our results align with studies showing that mistrust of benefits and concerns over side-effects are primary drivers of hesitancy. For example, a UK study of 32,361 adults identified these concerns as key drivers^19^, while a cross-sectional online survey of 492 participants in Ghana similarly linked vaccine mistrust and concerns about future effects to higher hesitancy on the Oxford COVID-19 Vaccine Hesitancy Scale (a 1–35 point measure), additionally noting lower hesitancy among single individuals^18^.

Our LCA approach provides a more granular demographic mapping than previous models. Paul et al. (2021)^19^ and Stead et al. (2021) ^20^ identified general “Uncertain” groups comprising 23% and 18.9% of their samples, respectively (with Paul et al. (2021) ^19^ noting women were more likely to be unsure or unwilling). Our study splits this demographic into the “Concerned Uptakers” (17.5%) and the “Undecided/Uninformed” (6.0%). Furthermore, we uniquely identified the “Unconcerned Refusers” (20.8%), who believe vaccines are effective but lack personal motivation. Methodologically, our identification of five classes aligns with other LCAs that found between three and nine classes^22–26^. As noted in previous literature, class membership is largely determined by formal education and age, and our five classes could theoretically be collapsed into three broader categories such as in Lamuda et al. (2023)^24^ (e.g., “High Trust,” “High Distrust,” and “Undecided”).

### 4.2. Strengths and Limitations

A key strength of using LCA is that it is multidimensional, able to identify distinct “profiles” of people based on their complex mix of responses. This provides a more nuanced understanding of vaccine attitude configurations than just looking at single population-wide averages. Furthermore, the inclusion of a broad range of sociodemographic factors allows a deeper understanding of who belongs to these profiles.

However, there are several limitations. As a secondary analysis, we had no influence on the specific questions asked; the survey items used inherently shape the identified classes, and different questions might yield different structures. Furthermore, important variables may not have been collected, leaving unmeasured confounders (e.g., detailed health literacy, specific sources of misinformation exposure, and psychological traits) that could influence class membership. Although we used multiple fit indices and prioritized interpretability, the choice of the number of classes in LCA still involves some subjectivity. Finally, although London has a diverse population, our findings may not be broadly generalizable to other UK cities, rural areas, or internationally due to differing socio-cultural contexts and healthcare systems.

## 5. Conclusion

This study identified five distinct latent classes of vaccine attitudes in a London-based community sample, revealing significant heterogeneity in how individuals perceive vaccines. The existence of these distinct belief profiles strongly suggests that universal public health campaigns are unlikely to be effective for all segments of the population. Public health campaigns require tailored messaging that clearly communicates benefits versus risks and addresses specific uncertainties. Furthermore, reinforcing the positive views of compliant groups and encouraging them to act as advocates within their networks could prove highly beneficial.

Future qualitative research should further explore the nuances of belief within each identified class. In Addition, longitudinal studies are needed to track how class membership changes over time or in response to public health campaigns and major events. Ultimately, future research could test the efficacy of interventions tailored specifically to the profiles of these latent classes.

Ultimately, this study demonstrates that moving beyond a rigid ‘Pro’ or ‘Anti’ vaccination binary framework is vital for contemporary public health understanding. As an ongoing initiative, the operational choices, resource distributions, and policy directions of the *Why We Get Vaccinated* campaign will directly benefit from these profiles.

## Data Availability

All data produced in the present study are available upon reasonable request to the authors

