## Supplemental Figures and Tables for "Moving Beyond the ‘Pro’ or ‘Anti’ Binary: Insights into Vaccination Attitudes and Behaviour, with a Case Study from a London, UK Survey"

---

### 1. Summary tables

| Characteristic | Don't know/ can't recall ($N = 95$) | No, I didn't ($N = 1006$) | Yes, I did ($N = 952$) |
|---|---|---|---|
| **Age** | | | |
| 18–24 | 29 (30·5%) | 122 (12·1%) | 91 (9·6%) |
| 25–34 | 36 (37·9%) | 353 (35·1%) | 192 (20·2%) |
| 35–44 | 13 (13·7%) | 180 (17·9%) | 138 (14·5%) |
| 45–54 | 9 (9·5%) | 156 (15·5%) | 96 (10·1%) |
| $\geq 55$ | 8 (8·4%) | 195 (19·4%) | 435 (45·7%) |
| **Gender** | | | |
| Female | 53 (55·8%) | 516 (51·3%) | 458 (48·1%) |
| Male | 42 (44·2%) | 490 (48·7%) | 494 (51·9%) |
| **Social Grade** | | | |
| High | 53 (55·8%) | 612 (60·8%) | 593 (62·3%) |
| Low | 42 (44·2%) | 394 (39·2%) | 359 (37·7%) |
| **Ethnicity** | | | |
| White | 43 (45·3%) | 612 (60·8%) | 699 (73·4%) |
| Asian | 6 (6·3%) | 140 (13·9%) | 83 (8·7%) |
| Black | 20 (21·1%) | 119 (11·8%) | 69 (7·2%) |
| Mixed | 20 (21·1%) | 103 (10·2%) | 82 (8·6%) |
| Other | 0 (0·0%) | 14 (1·4%) | 9 (0·9%) |
| Prefer not to say | 6 (6·3%) | 18 (1·8%) | 10 (1·1%) |
| **Marital Status** | | | |
| Married/ Civil Partnership | 27 (28·4%) | 323 (32·1%) | 413 (43·4%) |
| Living as married | 5 (5·3%) | 90 (8·9%) | 69 (7·2%) |
| Never Married | 52 (54·7%) | 492 (48·9%) | 339 (35·6%) |
| Separated/ Divorced | 4 (4·2%) | 54 (5·4%) | 73 (7·7%) |
| Widowed | 2 (2·1%) | 18 (1·8%) | 51 (5·4%) |
| Missing | 5 (5·3%) | 29 (2·9%) | 7 (0·7%) |
| **Housing Tenure** | | | |
| Own | 19 (20·0%) | 358 (35·6%) | 532 (55·9%) |
| Rent-free | 17 (17·9%) | 80 (8·0%) | 50 (5·3%) |
| Rent housing association | 4 (4·2%) | 70 (7·0%) | 55 (5·8%) |
| Rent private landlord | 23 (24·2%) | 264 (26·2%) | 150 (15·8%) |
| Rent local authority | 4 (4·2%) | 61 (6·1%) | 53 (5·6%) |
| Family/friends pay rent | 7 (7·4%) | 59 (5·9%) | 31 (3·3%) |
| Other | 11 (11·6%) | 35 (3·5%) | 19 (2·0%) |
| Missing | 10 (10·5%) | 79 (7·9%) | 62 (6·5%) |
| **Working Status** | | | |
| Working full-time | 39 (41·1%) | 554 (55·1%) | 417 (43·8%) |
| Working part-time | 15 (15·8%) | 155 (15·4%) | 135 (14·2%) |
| Full-time student | 16 (16·8%) | 57 (5·7%) | 31 (3·3%) |
| Not working/ Other | 17 (17·9%) | 91 (9·0%) | 75 (7·9%) |
| Retired | 2 (2·1%) | 68 (6·8%) | 258 (27·1%) |
| Unemployed | 6 (6·3%) | 81 (8·1%) | 36 (3·8%) |
| **Parent/Guardian of Child Aged** | | | |
| 4 years and under | 10 (10·5%) | 70 (7·0%) | 86 (9·0%) |
| 5 to 11 years | 7 (7·4%) | 138 (13·7%) | 72 (7·6%) |
| 12 to 16 years | 6 (6·3%) | 64 (6·4%) | 52 (5·5%) |
| 17 to 18 years | 2 (2·1%) | 17 (1·7%) | 13 (1·4%) |
| Over 18 years | 3 (3·2%) | 137 (13·6%) | 270 (28·4%) |
| Not parent/ guardian | 67 (70·5%) | 580 (57·7%) | 459 (48·2%) |
| **Number of Children in Household** | | | |
| 0 | 36 (37·9%) | 615 (61·1%) | 646 (67·9%) |
| 1 | 14 (14·7%) | 139 (13·8%) | 116 (12·2%) |
| 2 | 9 (9·5%) | 140 (13·9%) | 87 (9·1%) |
| $\geq 3$ | 8 (8·4%) | 58 (5·8%) | 58 (6·1%) |
| Refused | 28 (29·5%) | 54 (5·4%) | 45 (4·7%) |



Table 1: Demographic counts and percentage statistics from the WWGV survey by answer to question "Did you get the flu vaccine last winter?".

| Characteristic | Don't know/ unsure ($N = 176$) | Fairly likely ($N = 403$) | Not at all likely ($N = 435$) | Not very likely ($N = 303$) | Very likely ($N = 736$) |
|---|---|---|---|---|---|
| **Age** | | | | | |
| 18–24 | 32 (18·2%) | 86 (21·3%) | 42 (9·7%) | 38 (12·5%) | 44 (6·0%) |
| 25–34 | 76 (43·2%) | 146 (36·2%) | 122 (28·0%) | 107 (35·3%) | 130 (17·7%) |
| 35–44 | 19 (10·8%) | 62 (15·4%) | 95 (21·8%) | 54 (17·8%) | 101 (13·7%) |
| 45–54 | 21 (11·9%) | 38 (9·4%) | 74 (17·0%) | 51 (16·8%) | 77 (10·5%) |
| $\geq 55$ | 28 (15·9%) | 71 (17·6%) | 102 (23·4%) | 53 (17·5%) | 384 (52·2%) |
| **Gender** | | | | | |
| Female | 75 (42·6%) | 181 (44·9%) | 249 (57·2%) | 155 (51·2%) | 367 (49·9%) |
| Male | 101 (57·4%) | 222 (55·1%) | 186 (42·8%) | 148 (48·8%) | 369 (50·1%) |
| **Social Grade** | | | | | |
| High | 110 (62·5%) | 253 (62·8%) | 231 (53·1%) | 192 (63·4%) | 472 (64·1%) |
| Low | 66 (37·5%) | 150 (37·2%) | 204 (46·9%) | 111 (36·6%) | 264 (35·9%) |
| **Ethnicity** | | | | | |
| White | 110 (62·5%) | 227 (56·3%) | 267 (61·4%) | 189 (62·4%) | 561 (76·2%) |
| Asian | 21 (11·9%) | 46 (11·4%) | 64 (14·7%) | 41 (13·5%) | 57 (7·7%) |
| Black | 16 (9·1%) | 52 (12·9%) | 55 (12·6%) | 32 (10·6%) | 53 (7·2%) |
| Mixed | 21 (11·9%) | 66 (16·4%) | 36 (8·3%) | 28 (9·2%) | 54 (7·3%) |
| Other | 2 (1·1%) | 4 (1·0%) | 6 (1·4%) | 5 (1·7%) | 6 (0·8%) |
| Prefer not to say | 6 (3·4%) | 8 (2·0%) | 7 (1·6%) | 8 (2·6%) | 5 (0·7%) |
| **Marital Status** | | | | | |
| Married/ Civil Partnership | 50 (28·4%) | 144 (35·7%) | 122 (28·0%) | 106 (35·0%) | 341 (46·3%) |
| Living as married | 11 (6·2%) | 25 (6·2%) | 54 (12·4%) | 22 (7·3%) | 52 (7·1%) |
| Never Married | 85 (48·3%) | 206 (51·1%) | 218 (50·1%) | 144 (47·5%) | 230 (31·2%) |
| Separated/ Divorced | 6 (3·4%) | 16 (4·0%) | 24 (5·5%) | 21 (6·9%) | 64 (8·7%) |
| Widowed | 5 (2·8%) | 8 (2·0%) | 12 (2·8%) | 2 (0·7%) | 44 (6·0%) |
| Missing | 19 (10·8%) | 4 (1·0%) | 5 (1·1%) | 8 (2·6%) | 5 (0·7%) |
| **Housing Tenure** | | | | | |
| Own | 70 (39·8%) | 143 (35·5%) | 136 (31·3%) | 118 (38·9%) | 442 (60·1%) |
| Rent-free | 17 (9·7%) | 42 (10·4%) | 33 (7·6%) | 24 (7·9%) | 31 (4·2%) |
| Rent housing association | 8 (4·5%) | 25 (6·2%) | 40 (9·2%) | 15 (5·0%) | 41 (5·6%) |
| Rent private landlord | 35 (19·9%) | 88 (21·8%) | 131 (30·1%) | 76 (25·1%) | 107 (14·5%) |
| Rent local authority | 7 (4·0%) | 34 (8·4%) | 27 (6·2%) | 11 (3·6%) | 39 (5·3%) |
| Family/ friends pay rent | 10 (5·7%) | 26 (6·5%) | 24 (5·5%) | 16 (5·3%) | 21 (2·9%) |
| Other | 14 (8·0%) | 15 (3·7%) | 11 (2·5%) | 13 (4·3%) | 12 (1·6%) |
| Missing | 15 (8·5%) | 30 (7·4%) | 33 (7·6%) | 30 (9·9%) | 43 (5·8%) |
| **Working Status** | | | | | |
| Working full-time | 99 (56·2%) | 206 (51·1%) | 227 (52·2%) | 162 (53·5%) | 316 (42·9%) |
| Working part-time | 19 (10·8%) | 76 (18·9%) | 66 (15·2%) | 55 (18·2%) | 89 (12·1%) |
| Full-time student | 18 (10·2%) | 30 (7·4%) | 19 (4·4%) | 21 (6·9%) | 16 (2·2%) |
| Not working/ Other | 20 (11·4%) | 39 (9·7%) | 47 (10·8%) | 25 (8·3%) | 52 (7·1%) |
| Retired | 9 (5·1%) | 26 (6·5%) | 42 (9·7%) | 17 (5·6%) | 234 (31·8%) |
| Unemployed | 11 (6·2%) | 26 (6·5%) | 34 (7·8%) | 23 (7·6%) | 29 (3·9%) |
| **Parent/Guardian of Child Aged** | | | | | |
| 4 years and under | 9 (5·1%) | 51 (12·7%) | 26 (6·0%) | 25 (8·3%) | 55 (7·5%) |
| 5 to 11 years | 45 (25·6%) | 50 (12·4%) | 35 (8·0%) | 32 (10·6%) | 55 (7·5%) |
| 12 to 16 years | 7 (4·0%) | 28 (6·9%) | 25 (5·7%) | 23 (7·6%) | 39 (5·3%) |
| 17 to 18 years | 3 (1·7%) | 10 (2·5%) | 5 (1·1%) | 6 (2·0%) | 8 (1·1%) |
| Over 18 years | 16 (9·1%) | 45 (11·2%) | 73 (16·8%) | 34 (11·2%) | 242 (32·9%) |
| Not parent/ guardian | 96 (54·5%) | 219 (54·3%) | 271 (62·3%) | 183 (60·4%) | 337 (45·8%) |
| **Number of Children in Household** | | | | | |
| 0 | 77 (43·8%) | 192 (47·6%) | 305 (70·1%) | 186 (61·4%) | 537 (73·0%) |
| 1 | 25 (14·2%) | 64 (15·9%) | 51 (11·7%) | 47 (15·5%) | 82 (11·1%) |
| 2 | 45 (25·6%) | 57 (14·1%) | 39 (9·0%) | 33 (10·9%) | 62 (8·4%) |
| $\geq 3$ | 9 (5·1%) | 51 (12·7%) | 16 (3·7%) | 18 (5·9%) | 30 (4·1%) |
| Refused | 20 (11·4%) | 39 (9·7%) | 24 (5·5%) | 19 (6·3%) | 25 (3·4%) |

Table 2: Demographic counts and percentage statistics from the WWGV survey by answer to question "How likely, if at all, are you to get the flu vaccine this year?".

| Characteristic | Don't know/ unsure ($N = 67$) | No, I haven't ($N = 95$) | Not applicable ($N = 51$) | Not parent ($N = 1106$) | Yes, I have ($N = 734$) |
|---|---|---|---|---|---|
| **Age** | | | | | |
| 18–24 | 4 (6·0%) | 9 (9·5%) | 2 (3·9%) | 197 (17·8%) | 30 (4·1%) |
| 25–34 | 10 (14·9%) | 24 (25·3%) | 20 (39·2%) | 383 (34·6%) | 144 (19·6%) |
| 35–44 | 8 (11·9%) | 15 (15·8%) | 5 (9·8%) | 177 (16·0%) | 126 (17·2%) |
| 45–54 | 10 (14·9%) | 13 (13·7%) | 2 (3·9%) | 115 (10·4%) | 121 (16·5%) |
| $\geq$ 55 | 35 (52·2%) | 34 (35·8%) | 22 (43·1%) | 234 (21·2%) | 313 (42·6%) |
| **Gender** | | | | | |
| Female | 30 (44·8%) | 44 (46·3%) | 19 (37·3%) | 565 (51·1%) | 369 (50·3%) |
| Male | 37 (55·2%) | 51 (53·7%) | 32 (62·7%) | 541 (48·9%) | 365 (49·7%) |
| **Social Grade** | | | | | |
| High | 32 (47·8%) | 57 (60·0%) | 31 (60·8%) | 658 (59·5%) | 480 (65·4%) |
| Low | 35 (52·2%) | 38 (40·0%) | 20 (39·2%) | 448 (40·5%) | 254 (34·6%) |
| **Ethnicity** | | | | | |
| White | 46 (68·7%) | 63 (66·3%) | 27 (52·9%) | 688 (62·2%) | 530 (72·2%) |
| Asian | 9 (13·4%) | 6 (6·3%) | 2 (3·9%) | 132 (11·9%) | 80 (10·9%) |
| Black | 5 (7·5%) | 15 (15·8%) | 15 (29·4%) | 107 (9·7%) | 66 (9·0%) |
| Mixed | 4 (6·0%) | 10 (10·5%) | 6 (11·8%) | 141 (12·7%) | 44 (6·0%) |
| Other | 1 (1·5%) | 0 (0·0%) | 1 (2·0%) | 14 (1·3%) | 7 (1·0%) |
| Prefer not to say | 2 (3·0%) | 1 (1·1%) | 0 (0·0%) | 24 (2·2%) | 7 (1·0%) |
| **Marital Status** | | | | | |
| Married/ Civil Partnership | 41 (61·2%) | 55 (57·9%) | 19 (37·3%) | 159 (14·4%) | 489 (66·6%) |
| Living as married | 0 (0·0%) | 4 (4·2%) | 4 (7·8%) | 113 (10·2%) | 43 (5·9%) |
| Never Married | 10 (14·9%) | 21 (22·1%) | 18 (35·3%) | 751 (67·9%) | 83 (11·3%) |
| Separated/ Divorced | 12 (17·9%) | 10 (10·5%) | 5 (9·8%) | 39 (3·5%) | 65 (8·9%) |
| Widowed | 3 (4·5%) | 5 (5·3%) | 4 (7·8%) | 25 (2·3%) | 34 (4·6%) |
| Missing | 1 (1·5%) | 0 (0·0%) | 1 (2·0%) | 19 (1·7%) | 20 (2·7%) |
| **Housing Tenure** | | | | | |
| Own | 39 (58·2%) | 35 (36·8%) | 17 (33·3%) | 354 (32·0%) | 464 (63·2%) |
| Rent-free | 2 (3·0%) | 4 (4·2%) | 1 (2·0%) | 133 (12·0%) | 7 (1·0%) |
| Rent housing association | 8 (11·9%) | 11 (11·6%) | 4 (7·8%) | 55 (5·0%) | 51 (7·0%) |
| Rent private landlord | 11 (16·4%) | 25 (26·3%) | 6 (11·8%) | 294 (26·6%) | 101 (13·8%) |
| Rent local authority | 2 (3·0%) | 12 (12·6%) | 16 (31·4%) | 42 (3·8%) | 46 (6·3%) |
| Family/ friends pay rent | 1 (1·5%) | 2 (2·1%) | 1 (2·0%) | 83 (7·5%) | 10 (1·4%) |
| Other | 0 (0·0%) | 2 (2·1%) | 3 (5·9%) | 44 (4·0%) | 16 (2·2%) |
| Missing | 4 (6·0%) | 4 (4·2%) | 3 (5·9%) | 101 (9·1%) | 39 (5·3%) |
| **Working Status** | | | | | |
| Working full-time | 30 (44·8%) | 39 (41·1%) | 21 (41·2%) | 556 (50·3%) | 364 (49·6%) |
| Working part-time | 4 (6·0%) | 24 (25·3%) | 7 (13·7%) | 140 (12·7%) | 130 (17·7%) |
| Full-time student | 0 (0·0%) | 2 (2·1%) | 1 (2·0%) | 100 (9·0%) | 1 (0·1%) |
| Not working/ Other | 6 (9·0%) | 7 (7·4%) | 4 (7·8%) | 108 (9·8%) | 58 (7·9%) |
| Retired | 25 (37·3%) | 20 (21·1%) | 15 (29·4%) | 105 (9·5%) | 163 (22·2%) |
| Unemployed | 2 (3·0%) | 3 (3·2%) | 3 (5·9%) | 97 (8·8%) | 18 (2·5%) |
| **Parent/Guardian of Child Aged** | | | | | |
| 4 years and under | 9 (13·4%) | 18 (18·9%) | 10 (19·6%) | 0 (0·0%) | 129 (17·6%) |
| 5 to 11 years | 10 (14·9%) | 24 (25·3%) | 15 (29·4%) | 0 (0·0%) | 168 (22·9%) |
| 12 to 16 years | 8 (11·9%) | 14 (14·7%) | 1 (2·0%) | 0 (0·0%) | 99 (13·5%) |
| 17 to 18 years | 3 (4·5%) | 4 (4·2%) | 0 (0·0%) | 0 (0·0%) | 25 (3·4%) |
| Over 18 years | 37 (55·2%) | 35 (36·8%) | 25 (49·0%) | 0 (0·0%) | 313 (42·6%) |
| Not parent/ guardian | 0 (0·0%) | 0 (0·0%) | 0 (0·0%) | 1106 (100·0%) | 0 (0·0%) |
| **Number of Children in Household** | | | | | |
| 0 | 36 (53·7%) | 38 (40·0%) | 24 (47·1%) | 879 (79·5%) | 320 (43·6%) |
| 1 | 11 (16·4%) | 28 (29·5%) | 8 (15·7%) | 66 (6·0%) | 156 (21·3%) |
| 2 | 9 (13·4%) | 14 (14·7%) | 16 (31·4%) | 27 (2·4%) | 170 (23·2%) |
| $\geq$ 3 | 5 (7·5%) | 10 (10·5%) | 0 (0·0%) | 41 (3·7%) | 68 (9·3%) |
| Refused | 6 (9·0%) | 5 (5·3%) | 3 (5·9%) | 93 (8·4%) | 20 (2·7%) |



Table 3: Demographic counts and percentage statistics from the WWGV survey by answer to question "Have you vaccinated your oldest child with one or more doses of the MMR vaccine?".

| Characteristic | Neither agree/disagree ($N = 228$) | Strongly agree ($N = 1090$) | Strongly disagree ($N = 20$) | Tend to agree ($N = 678$) | Tend to disagree ($N = 37$) |
|---|---|---|---|---|---|
| **Age** | | | | | |
| 18–24 | 29 (13·0%) | 107 (9·8%) | 4 (20·0%) | 99 (15·0%) | 3 (8·1%) |
| 25–34 | 77 (34·0%) | 306 (28·0%) | 3 (15·0%) | 189 (28·0%) | 6 (16·0%) |
| 35–44 | 38 (17·0%) | 187 (17·0%) | 1 (5·0%) | 94 (14·0%) | 11 (30·0%) |
| 45–54 | 39 (17·0%) | 121 (11·0%) | 2 (10·0%) | 93 (14·0%) | 6 (16·0%) |
| $\geq 55$ | 45 (20·0%) | 369 (34·0%) | 10 (50·0%) | 203 (30·0%) | 11 (30·0%) |
| **Gender** | | | | | |
| Female | 112 (49·0%) | 550 (50·0%) | 12 (60·0%) | 327 (48·0%) | 26 (70·0%) |
| Male | 116 (51·0%) | 540 (50·0%) | 8 (40·0%) | 351 (52·0%) | 11 (30·0%) |
| **Social Grade** | | | | | |
| High | 96 (42·0%) | 747 (69·0%) | 10 (50·0%) | 387 (57·0%) | 18 (49·0%) |
| Low | 132 (58·0%) | 343 (31·0%) | 10 (50·0%) | 291 (43·0%) | 19 (51·0%) |
| **Ethnicity** | | | | | |
| White | 103 (45·0%) | 799 (73·0%) | 12 (60·0%) | 415 (61·0%) | 25 (68·0%) |
| Asian | 44 (19·0%) | 97 (8·9%) | 3 (15·0%) | 82 (12·0%) | 3 (8·1%) |
| Black | 43 (19·0%) | 82 (7·5%) | 1 (5·0%) | 76 (11·0%) | 6 (16·%) |
| Mixed | 23 (10·0%) | 95 (8·7%) | 4 (20·0%) | 81 (12·0%) | 2 (5·4%) |
| Other | 7 (3·1%) | 6 (0·6%) | 0 (0·0%) | 9 (1·3%) | 1 (2·7%) |
| Prefer not to say | 8 (3·5%) | 11 (1·0%) | 0 (0·0%) | 15 (2·2%) | 0 (0·0%) |
| **Marital Status** | | | | | |
| Married/ Civil Partnership | 58 (25·0%) | 451 (41·0%) | 6 (30·0%) | 237 (35·0%) | 11 (30·0%) |
| Living as married | 14 (6·1%) | 95 (8·7%) | 1 (5·0%) | 50 (7·4%) | 4 (11·0%) |
| Never Married | 119 (52·0%) | 427 (39·0%) | 10 (50·0%) | 315 (46·0%) | 12 (32·0%) |
| Separated/ Divorced | 19 (8·3%) | 65 (6·0%) | 2 (10·0%) | 40 (5·9%) | 5 (14·0%) |
| Widowed | 7 (3·1%) | 39 (3·6%) | 1 (5·0%) | 20 (2·9%) | 4 (11·0%) |
| Missing | 11 (4·8%) | 13 (1·2%) | 0 (0·0%) | 16 (2·4%) | 1 (2·7%) |
| **Housing Tenure** | | | | | |
| Own | 65 (27·9%) | 543 (49·8%) | 7 (35·0%) | 286 (41·8%) | 8 (21·4%) |
| Rent-free | 22 (9·6%) | 66 (6·1%) | 3 (15·0%) | 55 (8·1%) | 1 (2·7%) |
| Rent housing assoc· | 15 (6·6%) | 57 (5·2%) | 2 (10·0%) | 46 (6·8%) | 9 (24·0%) |
| Rent private landlord | 58 (25·0%) | 224 (21·0%) | 4 (20·0%) | 143 (21·0%) | 8 (22·0%) |
| Rent local authority | 22 (9·6%) | 38 (3·5%) | 1 (5·0%) | 50 (7·4%) | 7 (19·0%) |
| Family/ friends pay rent | 11 (4·8%) | 52 (4·8%) | 2 (10·0%) | 31 (4·6%) | 1 (2·7%) |
| Other | 16 (7·0%) | 23 (2·1%) | 0 (0·0%) | 25 (3·7%) | 1 (2·7%) |
| Missing | 19 (8·3%) | 87 (8·0%) | 1 (5·0%) | 42 (6·2%) | 2 (5·4%) |
| **Working Status** | | | | | |
| Working full-time | 111 (49·0%) | 549 (50·0%) | 7 (35·0%) | 324 (48·0%) | 19 (51·0%) |
| Working part-time | 36 (16·0%) | 155 (14·0%) | 1 (5·0%) | 111 (16·0%) | 2 (5·4%) |
| Full-time student | 13 (5·7%) | 43 (3·9%) | 1 (5·0%) | 47 (6·9%) | 0 (0·0%) |
| Not working/ Other | 30 (13·0%) | 81 (7·4%) | 2 (10·0%) | 66 (9·7%) | 4 (11·0%) |
| Retired | 21 (9·2%) | 201 (18·0%) | 8 (40·0%) | 90 (13·0%) | 8 (22·0%) |
| Unemployed | 17 (7·5%) | 61 (5·6%) | 1 (5·0%) | 40 (5·9%) | 4 (11·0%) |
| **Parent/Guardian of Child Aged** | | | | | |
| 4 years and under | 15 (6·6%) | 96 (8·8%) | 2 (10·0%) | 47 (6·9%) | 6 (16·0%) |
| 5 to 11 years | 19 (8·3%) | 106 (9·7%) | 0 (0·0%) | 87 (13·0%) | 5 (14·0%) |
| 12 to 16 years | 14 (6·1%) | 62 (5·7%) | 0 (0·0%) | 45 (6·6%) | 1 (2·7%) |
| 17 to 18 years | 2 (0·9%) | 16 (1·5%) | 0 (0·0%) | 13 (1·9%) | 1 (2·7%) |
| Over 18 years | 29 (13·0%) | 237 (22·0%) | 5 (25·0%) | 129 (19·0%) | 10 (27·0%) |
| Not parent/ guardian | 149 (65·0%) | 573 (53·0%) | 13 (65·0%) | 357 (53·0%) | 14 (38·0%) |
| **Number of Children in Household** | | | | | |
| 0 | 135 (59·0%) | 726 (67·0%) | 16 (80·0%) | 398 (59·0%) | 22 (59·0%) |
| 1 | 33 (14·0%) | 140 (13·0%) | 2 (10·0%) | 89 (13·0%) | 5 (14·0%) |
| 2 | 15 (6·6%) | 127 (12·0%) | 1 (5·0%) | 88 (13·0%) | 5 (14·0%) |
| $\geq 3$ | 14 (6·1%) | 55 (5·0%) | 0 (0·0%) | 54 (8·0%) | 1 (2·7%) |
| Refused | 31 (14·0%) | 42 (3·9%) | 1 (5·0%) | 49 (7·2%) | 4 (11·0%) |

Table 4: Demographic counts and percentage statistics from the WWGV survey by answer to statement "Vaccines are effective".

| Characteristic | Neither agree/disagree ($N = 228$) | Strongly agree ($N = 1090$) | Strongly disagree ($N = 20$) | Tend to agree ($N = 678$) | Tend to disagree ($N = 37$) |
|---|---|---|---|---|---|
| **Age** | | | | | |
| 18–24 | 29 (12·7%) | 107 (9·8%) | 4 (20·0%) | 99 (14·6%) | 3 (8·1%) |
| 25–34 | 77 (33·8%) | 306 (28·1%) | 3 (15·0%) | 189 (27·9%) | 6 (16·2%) |
| 35–44 | 38 (16·7%) | 187 (17·2%) | 1 (5·0%) | 94 (13·9%) | 11 (29·7%) |
| 45–54 | 39 (17·1%) | 121 (11·1%) | 2 (10·0%) | 93 (13·7%) | 6 (16·2%) |
| $\geq$ 55 | 45 (19·7%) | 369 (33·9%) | 10 (50·0%) | 203 (29·9%) | 11 (29·7%) |
| **Gender** | | | | | |
| Female | 112 (49·1%) | 550 (50·5%) | 12 (60·0%) | 327 (48·2%) | 26 (70·3%) |
| Male | 116 (50·9%) | 540 (49·5%) | 8 (40·0%) | 351 (51·8%) | 11 (29·7%) |
| **Social Grade** | | | | | |
| High | 96 (42·1%) | 747 (68·5%) | 10 (50·0%) | 387 (57·1%) | 18 (48·6%) |
| Low | 132 (57·9%) | 343 (31·5%) | 10 (50·0%) | 291 (42·9%) | 19 (51·4%) |
| **Ethnicity** | | | | | |
| White | 103 (45·2%) | 799 (73·3%) | 12 (60·0%) | 415 (61·2%) | 25 (67·6%) |
| Asian | 44 (19·3%) | 97 (8·9%) | 3 (15·0%) | 82 (12·1%) | 3 (8·1%) |
| Black | 43 (18·9%) | 82 (7·5%) | 1 (5·0%) | 76 (11·2%) | 6 (16·2%) |
| Mixed | 23 (10·1%) | 95 (8·7%) | 4 (20·0%) | 81 (11·9%) | 2 (5·4%) |
| Other | 7 (3·1%) | 6 (0·6%) | 0 (0·0%) | 9 (1·3%) | 1 (2·7%) |
| Prefer not to say | 8 (3·5%) | 11 (1·0%) | 0 (0·0%) | 15 (2·2%) | 0 (0·0%) |
| **Marital Status** | | | | | |
| Married/ Civil Partnership | 58 (25·4%) | 451 (41·4%) | 6 (30·0%) | 237 (35·0%) | 11 (29·7%) |
| Living as married | 14 (6·1%) | 95 (8·7%) | 1 (5·0%) | 50 (7·4%) | 4 (10·8%) |
| Never Married | 119 (52·2%) | 427 (39·2%) | 10 (50·0%) | 315 (46·5%) | 12 (32·4%) |
| Separated/ Divorced | 19 (8·3%) | 65 (6·0%) | 2 (10·0%) | 40 (5·9%) | 5 (13·5%) |
| Widowed | 7 (3·1%) | 39 (3·6%) | 1 (5·0%) | 20 (2·9%) | 4 (10·8%) |
| Missing | 11 (4·8%) | 13 (1·2%) | 0 (0·0%) | 16 (2·4%) | 1 (2·7%) |
| **Housing Tenure** | | | | | |
| Own | 65 (28·5%) | 543 (49·8%) | 7 (35·0%) | 286 (42·2%) | 8 (21·6%) |
| Rent-free | 22 (9·6%) | 66 (6·1%) | 3 (15·0%) | 55 (8·1%) | 1 (2·7%) |
| Rent housing association | 15 (6·6%) | 57 (5·2%) | 2 (10·0%) | 46 (6·8%) | 9 (24·3%) |
| Rent private landlord | 58 (25·4%) | 224 (20·6%) | 4 (20·0%) | 143 (21·1%) | 8 (21·6%) |
| Rent local authority | 22 (9·6%) | 38 (3·5%) | 1 (5·0%) | 50 (7·4%) | 7 (18·9%) |
| Family/ friends pay rent | 11 (4·8%) | 52 (4·8%) | 2 (10·0%) | 31 (4·6%) | 1 (2·7%) |
| Other | 16 (7·0%) | 23 (2·1%) | 0 (0·0%) | 25 (3·7%) | 1 (2·7%) |
| Missing | 19 (8·3%) | 87 (8·0%) | 1 (5·0%) | 42 (6·2%) | 2 (5·4%) |
| **Working Status** | | | | | |
| Working full-time | 111 (48·7%) | 549 (50·4%) | 7 (35·0%) | 324 (47·8%) | 19 (51·4%) |
| Working part-time | 36 (15·8%) | 155 (14·2%) | 1 (5·0%) | 111 (16·4%) | 2 (5·4%) |
| Full-time student | 13 (5·7%) | 43 (3·9%) | 1 (5·0%) | 47 (6·9%) | 0 (0·0%) |
| Not working/ Other | 30 (13·2%) | 81 (7·4%) | 2 (10·0%) | 66 (9·7%) | 4 (10·8%) |
| Retired | 21 (9·2%) | 201 (18·4%) | 8 (40·0%) | 90 (13·3%) | 8 (21·6%) |
| Unemployed | 17 (7·5%) | 61 (5·6%) | 1 (5·0%) | 40 (5·9%) | 4 (10·8%) |
| **Parent/Guardian of Child Aged** | | | | | |
| 4 years and under | 15 (6·6%) | 96 (8·8%) | 2 (10·0%) | 47 (6·9%) | 6 (16·2%) |
| 5 to 11 years | 19 (8·3%) | 106 (9·7%) | 0 (0·0%) | 87 (12·8%) | 5 (13·5%) |
| 12 to 16 years | 14 (6·1%) | 62 (5·7%) | 0 (0·0%) | 45 (6·6%) | 1 (2·7%) |
| 17 to 18 years | 2 (0·9%) | 16 (1·5%) | 0 (0·0%) | 13 (1·9%) | 1 (2·7%) |
| Over 18 years | 29 (12·7%) | 237 (21·7%) | 5 (25·0%) | 129 (19·0%) | 10 (27·0%) |
| Not parent/ guardian | 149 (65·4%) | 573 (52·6%) | 13 (65·0%) | 357 (52·7%) | 14 (37·8%) |
| **Number of Children in Household** | | | | | |
| 0 | 135 (59·2%) | 726 (66·6%) | 16 (80·0%) | 398 (58·7%) | 22 (59·5%) |
| 1 | 33 (14·5%) | 140 (12·8%) | 2 (10·0%) | 89 (13·1%) | 5 (13·5%) |
| 2 | 15 (6·6%) | 127 (11·7%) | 1 (5·0%) | 88 (13·0%) | 5 (13·5%) |
| $\geq$ 3 | 14 (6·1%) | 55 (5·0%) | 0 (0·0%) | 54 (8·0%) | 1 (2·7%) |
| Refused | 31 (13·6%) | 42 (3·9%) | 1 (5·0%) | 49 (7·2%) | 4 (10·8%) |

Table 5: Demographic counts and percentage statistics from the WWGV survey by answer to statement “Vaccines are effective”.

| Characteristic | Neither agree/disagree ($N = 495$) | Strongly agree ($N = 329$) | Strongly disagree ($N = 267$) | Tend to agree ($N = 482$) | Tend to disagree ($N = 480$) |
|---|---|---|---|---|---|
| **Age** | | | | | |
| 18–24 | 53 (10·7%) | 56 (17·0%) | 14 (5·2%) | 87 (18·0%) | 32 (6·7%) |
| 25–34 | 136 (27·5%) | 124 (37·7%) | 47 (17·6%) | 157 (32·6%) | 117 (24·4%) |
| 35–44 | 75 (15·2%) | 59 (17·9%) | 43 (16·1%) | 66 (13·7%) | 88 (18·3%) |
| 45–54 | 73 (14·7%) | 30 (9·1%) | 36 (13·5%) | 63 (13·1%) | 59 (12·3%) |
| $\geq 55$ | 158 (31·9%) | 60 (18·2%) | 127 (47·6%) | 109 (22·6%) | 184 (38·3%) |
| **Gender** | | | | | |
| Female | 262 (52·9%) | 158 (48·0%) | 119 (44·6%) | 240 (49·8%) | 248 (51·7%) |
| Male | 233 (47·1%) | 171 (52·0%) | 148 (55·4%) | 242 (50·2%) | 232 (48·3%) |
| **Social Grade** | | | | | |
| High | 286 (57·8%) | 168 (51·1%) | 201 (75·3%) | 271 (56·2%) | 332 (69·2%) |
| Low | 209 (42·2%) | 161 (48·9%) | 66 (24·7%) | 211 (43·8%) | 148 (30·8%) |
| **Ethnicity** | | | | | |
| White | 293 (59·2%) | 180 (54·7%) | 226 (84·6%) | 274 (56·8%) | 381 (79·4%) |
| Asian | 64 (12·9%) | 39 (11·9%) | 22 (8·2%) | 62 (12·9%) | 42 (8·8%) |
| Black | 60 (12·1%) | 53 (16·1%) | 6 (2·2%) | 68 (14·1%) | 21 (4·4%) |
| Mixed | 55 (11·1%) | 44 (13·4%) | 10 (3·7%) | 65 (13·5%) | 31 (6·5%) |
| Other | 8 (1·6%) | 6 (1·8%) | 1 (0·4%) | 5 (1·0%) | 3 (0·6%) |
| Prefer not to say | 15 (3·0%) | 7 (2·1%) | 2 (0·7%) | 8 (1·7%) | 2 (0·4%) |
| **Marital Status** | | | | | |
| Married/ Civil Partnership | 176 (35·6%) | 118 (35·9%) | 119 (44·6%) | 156 (32·4%) | 194 (40·4%) |
| Living as married | 35 (7·1%) | 17 (5·2%) | 27 (10·1%) | 28 (5·8%) | 57 (11·9%) |
| Never Married | 208 (42·0%) | 158 (48·0%) | 97 (36·3%) | 249 (51·7%) | 171 (35·6%) |
| Separated/ Divorced | 39 (7·9%) | 19 (5·8%) | 13 (4·9%) | 27 (5·6%) | 33 (6·9%) |
| Widowed | 22 (4·4%) | 12 (3·6%) | 11 (4·1%) | 12 (2·5%) | 14 (2·9%) |
| Missing | 15 (3·0%) | 5 (1·5%) | 0 (0·0%) | 10 (2·1%) | 11 (2·3%) |
| **Housing Tenure** | | | | | |
| Own | 225 (45·5%) | 110 (33·4%) | 160 (60·0%) | 161 (33·4%) | 253 (52·7%) |
| Rent-free | 31 (6·3%) | 29 (8·8%) | 16 (6·0%) | 48 (10·0%) | 23 (4·8%) |
| Rent housing association | 35 (7·1%) | 21 (6·4%) | 6 (2·2%) | 37 (7·7%) | 30 (6·3%) |
| Rent private landlord | 92 (18·6%) | 84 (25·5%) | 55 (20·6%) | 110 (22·8%) | 96 (20·0%) |
| Rent local authority | 26 (5·3%) | 26 (7·9%) | 8 (3·0%) | 38 (7·9%) | 20 (4·2%) |
| Family/ friends pay rent | 27 (5·5%) | 17 (5·2%) | 8 (3·0%) | 27 (5·6%) | 18 (3·8%) |
| Other | 19 (3·8%) | 10 (3·0%) | 2 (0·7%) | 18 (3·7%) | 16 (3·3%) |
| Missing | 40 (8·1%) | 32 (9·7%) | 12 (4·5%) | 43 (8·9%) | 24 (5·0%) |
| **Working Status** | | | | | |
| Working full-time | 241 (48·7%) | 163 (49·5%) | 133 (49·8%) | 227 (47·1%) | 246 (51·3%) |
| Working part-time | 77 (15·6%) | 64 (19·5%) | 25 (9·4%) | 89 (18·5%) | 50 (10·4%) |
| Full-time student | 29 (5·9%) | 20 (6·1%) | 8 (3·0%) | 33 (6·8%) | 14 (2·9%) |
| Not working/ Other | 43 (8·7%) | 27 (8·2%) | 22 (8·2%) | 53 (11·0%) | 38 (7·9%) |
| Retired | 75 (15·2%) | 31 (9·4%) | 70 (26·2%) | 46 (9·5%) | 106 (22·1%) |
| Unemployed | 30 (6·1%) | 24 (7·3%) | 9 (3·4%) | 34 (7·1%) | 26 (5·4%) |
| **Parent/Guardian of Child Aged** | | | | | |
| 4 years and under | 35 (7·1%) | 40 (12·2%) | 14 (5·2%) | 51 (10·6%) | 26 (5·4%) |
| 5 to 11 years | 63 (12·7%) | 54 (16·4%) | 11 (4·1%) | 49 (10·2%) | 40 (8·3%) |
| 12 to 16 years | 28 (5·7%) | 24 (7·3%) | 10 (3·7%) | 34 (7·1%) | 26 (5·4%) |
| 17 to 18 years | 10 (2·0%) | 4 (1·2%) | 3 (1·1%) | 9 (1·9%) | 6 (1·3%) |
| Over 18 years | 97 (19·6%) | 47 (14·3%) | 75 (28·1%) | 77 (16·0%) | 114 (23·8%) |
| Not parent/ guardian | 262 (52·9%) | 160 (48·6%) | 154 (57·7%) | 262 (54·4%) | 268 (55·8%) |
| **Number of Children in Household** | | | | | |
| 0 | 307 (62·0%) | 151 (45·9%) | 215 (80·5%) | 261 (54·1%) | 363 (75·6%) |
| 1 | 63 (12·7%) | 66 (20·1%) | 21 (7·9%) | 65 (13·5%) | 54 (11·3%) |
| 2 | 54 (10·9%) | 47 (14·3%) | 22 (8·2%) | 66 (13·7%) | 47 (9·8%) |
| $\geq 3$ | 30 (6·1%) | 33 (10·0%) | 5 (1·9%) | 47 (9·8%) | 9 (1·9%) |
| Refused | 41 (8·3%) | 32 (9·7%) | 4 (1·5%) | 43 (8·9%) | 7 (1·5%) |

Table 6: Demographic counts and percentage statistics from the WWGV survey by answer to question "I am concerned about serious adverse effects (i.e. negative side effects or unwanted results) of vaccines".

### 2. Separate regression results

#### *2.1. Tables*

| Characteristic | Log OR | SD | 95% CI |
|---|---|---|---|
| (Intercept) | 0·637 | 0·333 | 0·082, 1·184 |
| **Age** (ref: 18–24) | | | |
| 25–34 | -0·398 | 0·206 | -0·740, -0·071 |
| 35–44 | -0·063 | 0·221 | -0·435, 0·295 |
| 45–54 | -0·429 | 0·240 | -0·826, -0·048 |
| $\geq$ 55 | 0·363 | 0·244 | -0·044, 0·767 |
| **Gender** (ref: Female) | | | |
| Male | 0·141 | 0·098 | -0·026, 0·308 |
| Social Grade (ref: High) | | | |
| Low | 0·131 | 0·109 | -0·044, 0·313 |
| **Ethnicity** (ref: White) | | | |
| Asian | -0·486 | 0·149 | -0·729, -0·251 |
| Black | -0·247 | 0·169 | -0·525, 0·011 |
| Mixed | 0·186 | 0·186 | -0·109, 0·482 |
| Other | -0·135 | 0·481 | -0·928, 0·624 |
| Prefer not to say | -0·457 | 0·450 | -1·229, 0·286 |
| **Marital Status** (ref: Married/ Civil Partnership) | | | |
| Living as married | -0·296 | 0·197 | -0·630, 0·042 |
| Never Married | -0·225 | 0·152 | -0·466, 0·020 |
| Separated/ Divorced | 0·074 | 0·207 | -0·265, 0·430 |
| Widowed | 0·255 | 0·325 | -0·273, 0·795 |
| Missing | -1·046 | 0·478 | -1·857, -0·306 |
| **Housing Tenure** (ref: Own) | | | |
| Rent-free | -0·393 | 0·237 | -0·776, -0·006 |
| Rent from a housing association | -0·645 | 0·208 | -1·004, -0·317 |
| Rent from a private landlord | -0·537 | 0·138 | -0·770, -0·310 |
| Rent from my local authority | -0·339 | 0·223 | -0·702, 0·017 |
| Family/ friends but pay rent | -0·638 | 0·259 | -1·073, -0·203 |
| Other | -0·758 | 0·314 | -1·278, -0·238 |
| Missing | -0·234 | 0·209 | -0·571, 0·112 |
| **Working Status** (ref: Working full time) | | | |
| Working part-time | -0·072 | 0·150 | -0·312, 0·168 |
| Full-time student | -0·332 | 0·289 | -0·818, 0·125 |
| Not working/ Other | 0·065 | 0·185 | -0·239, 0·373 |
| Retired | 0·888 | 0·204 | 0·557, 1·230 |
| Unemployed | -0·529 | 0·219 | -0·890, -0·176 |
| **Parent/Guardian of Child Aged** (ref: 4 years and under) | | | |
| 5 to 11 years | -0·782 | 0·231 | -1·155, -0·415 |
| 12 to 16 years | -0·437 | 0·271 | -0·882, -0·008 |
| 17 to 18 years | -0·879 | 0·456 | -1·617, -0·145 |
| Over 18 years | -0·510 | 0·274 | -0·943, -0·067 |
| Not parent/ guardian | -0·343 | 0·239 | -0·727, 0·051 |
| **Number of Children in Household** (ref: 0) | | | |
| 1 | 0·164 | 0·206 | -0·177, 0·517 |
| 2 | -0·159 | 0·243 | -0·537, 0·236 |
| $\geq$ 3 | 0·174 | 0·260 | -0·260, 0·593 |
| Refused | 0·319 | 0·242 | -0·077, 0·721 |

Abbreviations: CI = Credible Interval, OR = Odds Ratio, SD = Standard Deviation

Table 7: Posterior Log ORs by covariate for answer to the question “Did you get the flu vaccine last winter?”. Reference categories are given in brackets.

| Characteristic | OR | 95% CI |
|---|---|---|
| **Age** (ref: 18–24) | | |
| 25-34 | 0·67 | 0·48, 0·93 |
| 35-44 | 0·94 | 0·65, 1·34 |
| 45-54 | 0·65 | 0·44, 0·95 |
| $\geq$ 55 | 1·44 | 0·96, 2·15 |
| **Gender** (ref: Female) | | |
| Male | 1·15 | 0·97, 1·36 |
| **Social Grade** (ref: High) | | |
| Low | 1·14 | 0·96, 1·37 |
| **Ethnicity** (ref: White) | | |
| Asian | 0·62 | 0·48, 0·78 |
| Black | 0·78 | 0·59, 1·01 |
| Mixed | 1·20 | 0·90, 1·62 |
| Other | 0·87 | 0·40, 1·87 |
| Prefer not to say | 0·63 | 0·29, 1·33 |
| **Marital Status** (ref: Married/ Civil Partnership) | | |
| Living as married | 0·74 | 0·53, 1·04 |
| Never Married | 0·80 | 0·63, 1·12 |
| Separated/ Divorced | 1·08 | 0·77, 1·54 |
| Widowed | 1·29 | 0·76, 2·21 |
| Missing | 0·35 | 0·16, 0·74 |
| **Housing Tenure** (ref: Own) | | |
| Rent-free | 0·68 | 0·46, 0·99 |
| Rent from a housing association | 0·52 | 0·37, 0·73 |
| Rent from a private landlord | 0·58 | 0·46, 0·73 |
| Rent from my local authority | 0·71 | 0·50, 1·02 |
| Family/ friends but pay rent | 0·53 | 0·34, 0·82 |
| Other | 0·47 | 0·28, 0·79 |
| Missing | 0·79 | 0·56, 1·12 |
| **Working Status** (ref: Working full-time) | | |
| Working part-time | 0·93 | 0·73, 1·18 |
| Full-time student | 0·72 | 0·44, 1·13 |
| Not working/ Other | 1·07 | 0·79, 1·45 |
| Retired | 2·43 | 1·75, 3·42 |
| Unemployed | 0·59 | 0·41, 0·84 |
| **Parent/Guardian of Child Aged** (ref: 4 years and under) | | |
| 5 to 11 years | 0·46 | 0·32, 0·66 |
| 12 to 16 years | 0·65 | 0·41, 0·99 |
| 17 to 18 years | 0·42 | 0·20, 0·87 |
| Over 18 years | 0·60 | 0·39, 0·94 |
| Not parent/ guardian | 0·71 | 0·48, 1·05 |
| **Number of Children in Household** (ref: 0) | | |
| 1 | 1·18 | 0·84, 1·68 |
| 2 | 0·85 | 0·58, 1·27 |
| $\geq$ 3 | 1·19 | 0·77, 1·81 |
| Refused | 1·38 | 0·93, 2·06 |



Abbreviations: CI = Credible Interval, OR = Odds Ratio

Table 8: Posterior ORs by covariate for answer to the question “Did you get the flu vaccine last winter?”. Reference categories are given in brackets.

| **Characteristic** | **Log OR** | **SD** | **95% CI** |
|---|---|---|---|
| (Intercept) | 1·088 | 0·343 | 0·546, 1·672 |
| **Age** (ref: 18–24) | | | |
| 25–34 | -0·391 | 0·206 | -0·722, -0·055 |
| 35–44 | -0·548 | 0·215 | -0·919, -0·200 |
| 45–54 | -0·839 | 0·231 | -1·236, -0·458 |
| $\geq$ 55 | 0·040 | 0·242 | -0·372, 0·431 |
| **Gender** (ref: Female) | | | |
| Male | 0·244 | 0·108 | 0·080, 0·415 |
| **Social Grade** (ref: High) | | | |
| Low | -0·179 | 0·107 | -0·361, -0·001 |
| **Ethnicity** (ref: White) | | | |
| Asian | -0·313 | 0·143 | -0·561, -0·083 |
| Black | -0·027 | 0·156 | -0·296, 0·231 |
| Mixed | 0·445 | 0·186 | 0·150, 0·763 |
| Other | -0·172 | 0·493 | -0·972, 0·620 |
| Prefer not to say | -0·457 | 0·430 | -1·241, 0·173 |
| **Marital Status** (ref: Married/ Civil Partnership) | | | |
| Living as married | -0·396 | 0·200 | -0·724, -0·061 |
| Never Married | -0·182 | 0·144 | -0·427, 0·061 |
| Separated/ Divorced | 0·092 | 0·216 | -0·260, 0·452 |
| Widowed | 0·307 | 0·334 | -0·235, 0·908 |
| Missing | -0·831 | 0·478 | -1·652, -0·068 |
| **Housing Tenure** (ref: Own) | | | |
| Rent-free | -0·350 | 0·224 | -0·719, 0·017 |
| Rent from a housing association | -0·557 | 0·210 | -0·902, -0·208 |
| Rent from a private landlord | -0·482 | 0·142 | -0·711, -0·255 |
| Rent from my local authority | 0·019 | 0·230 | -0·358, 0·394 |
| Family/ friends but pay rent | -0·287 | 0·248 | -0·689, 0·132 |
| Other | -0·437 | 0·314 | -0·942, 0·080 |
| Missing | -0·437 | 0·216 | -0·776, -0·099 |
| **Working Status** (ref: Working full-time) | | | |
| Working part-time | -0·142 | 0·149 | -0·383, 0·110 |
| Full-time student | -0·386 | 0·267 | -0·840, 0·060 |
| Not working/ Other | -0·005 | 0·175 | -0·309, 0·282 |
| Retired | 0·627 | 0·205 | 0·282, 0·980 |
| Unemployed | -0·116 | 0·214 | -0·470, 0·229 |
| **Parent/Guardian of Child Aged** (ref: 4 years and under) | | | |
| 5 to 11 years | -0·231 | 0·227 | -0·615, 0·166 |
| 12 to 16 years | -0·154 | 0·274 | -0·592, 0·284 |
| 17 to 18 years | -0·355 | 0·461 | -1·083, 0·414 |
| Over 18 years | -0·197 | 0·305 | -0·657, 0·272 |
| Not parent/ guardian | -0·224 | 0·251 | -0·651, 0·180 |
| **Number of Children in Household** (ref: 0) | | | |
| 1 | 0·224 | 0·215 | -0·132, 0·572 |
| 2 | 0·118 | 0·243 | -0·285, 0·517 |
| $\geq$ 3 | 0·477 | 0·260 | 0·042, 0·897 |
| Refused | 0·302 | 0·231 | -0·066, 0·693 |

Abbreviations: CI = Credible Interval, OR = Odds Ratio, SD = Standard Deviation

Table 9: Posterior Log OR by covariate for answer to the question "How likely, if at all, are you to get the flu vaccine this year?".

| Characteristic | OR | 95% CI |
|---|---|---|
| **Age** (ref: 18–24) | | |
| 25–34 | 0·68 | 0·49, 0·95 |
| 35–44 | 0·58 | 0·40, 0·82 |
| 45–54 | 0·43 | 0·29, 0·63 |
| $\geq$ 55 | 1·04 | 0·69, 1·54 |
| **Gender** (ref: Female) | | |
| Male | 1·28 | 1·08, 1·51 |
| **Social Grade** (ref: High) | | |
| Low | 0·84 | 0·70, 1·00 |
| **Ethnicity** (ref: White) | | |
| Asian | 0·73 | 0·57, 0·92 |
| Black | 0·97 | 0·74, 1·26 |
| Mixed | 1·56 | 1·16, 2·14 |
| Other | 0·84 | 0·38, 1·86 |
| Prefer not to say | 0·58 | 0·29, 1·19 |
| **Marital Status** (ref: Married/ Civil Partnership) | | |
| Living as married | 0·67 | 0·48, 0·94 |
| Never Married | 0·83 | 0·65, 1·06 |
| Separated/ Divorced | 1·10 | 0·77, 1·57 |
| Widowed | 1·36 | 0·79, 2·48 |
| Missing | 0·44 | 0·19, 0·93 |
| **Housing Tenure** (ref: Own) | | |
| Rent-free | 0·70 | 0·49, 1·02 |
| Rent from a housing association | 0·57 | 0·41, 0·81 |
| Rent from a private landlord | 0·62 | 0·49, 0·77 |
| Rent from my local authority | 1·02 | 0·70, 1·148 |
| Family/ friends but pay rent | 0·75 | 0·50, 1·14 |
| Other | 0·65 | 0·39, 1·08 |
| Missing | 0·65 | 0·46, 0·91 |
| **Working Status** (ref: Working full-time) | | |
| Working part time | 0·87 | 0·68, 1·12 |
| Full time student | 0·68 | 0·43, 1·06 |
| Not working/ Other | 1·00 | 0·73, 1·33 |
| Retired | 1·87 | 1·33, 2·66 |
| Unemployed | 0·89 | 0·63, 1·26 |
| **Parent/Guardian of Child Aged** (ref: 4 years and under) | | |
| 5 to 11 years | 0·79 | 0·54, 1·18 |
| 12 to 16 years | 0·86 | 0·55, 1·33 |
| 17 to 18 years | 0·70 | 0·34, 1·51 |
| Over 18 years | 0·82 | 0·52, 1·31 |
| Not parent/ guardian | 0·80 | 0·52, 1·20 |
| **Number of Children in Household** (ref: 0) | | |
| 1 | 1·25 | 0·88, 1·77 |
| 2 | 1·13 | 0·75, 1·68 |
| $\geq$ 3 | 1·61 | 1·04, 2·45 |
| Refused | 1·35 | 0·94, 2·00 |



Abbreviations: CI = Credible Interval, OR = Odds Ratio

Table 10: Posterior OR by covariate for answer to question "How likely, if at all, are you to get the flu vaccine this year?".

| Characteristic | Log OR | SD | 95% CI |
|---|---|---|---|
| (Intercept) | 2·631 | 0·590 | 1·668, 3·600 |
| **Age** (ref: 18–24) | | | |
| 25–34 | -0·261 | 0·408 | -0·976, 0·435 |
| 35–44 | 0·233 | 0·437 | -0·493, 0·946 |
| 45–54 | 0·335 | 0·454 | -0·408, 1·071 |
| $\geq$ 55 | 0·544 | 0·499 | -0·277, 1·338 |
| **Gender** (ref: Female) | | | |
| Male | -0·462 | 0·244 | -0·853, -0·064 |
| **Social Grade** (ref: High) | | | |
| Low | -0·049 | 0·239 | -0·455, 0·350 |
| **Ethnicity** (ref: White) | | | |
| Asian | 0·573 | 0·392 | -0·026, 1·255 |
| Black | -0·306 | 0·307 | -0·820, 0·217 |
| Mixed | -0·048 | 0·429 | -0·718, 0·655 |
| Other | | | |
| Prefer not to say | 0·215 | 0·770 | -1·033, 1·566 |
| **Marital Status** (ref: Married/ Civil Partnership) | | | |
| Living as married | 0·164 | 0·484 | -0·569, 1·002 |
| Never Married | -0·306 | 0·323 | -0·804, 0·233 |
| Separated/ Divorced | -0·262 | 0·371 | -0·859, 0·388 |
| Widowed | -0·293 | 0·506 | -1·080, 0·571 |
| **Housing Tenure** (ref: Own outright) | | | |
| Own with a mortgage | | | |
| Rent-free | -1·146 | 0·661 | -2·250, -0·005 |
| Rent from a housing association | -0·757 | 0·392 | -1·387, -0·111 |
| Rent from a private landlord | -0·733 | 0·310 | -1·244, -0·231 |
| Rent from my local authority | -0·827 | 0·376 | -1·439, -0·182 |
| Family/ friends but pay rent | -0·422 | 0·640 | -1·395, 0·743 |
| Shared ownership scheme | | | |
| Other | -0·240 | 0·668 | -1·1312, 0·922 |
| Missing | 0·040 | 0·516 | -0·763, 0·908 |
| **Working Status** (ref: Working full time) | | | |
| Working part time | -0·309 | 0·301 | -0·820, 0·191 |
| Full time student | -1·177 | 0·814 | -2·481, 0·144 |
| Not working/ Other | 0·189 | 0·437 | -0·520, 0·923 |
| Retired | -0·399 | 0·385 | -1·041, 0·210 |
| Unemployed | 0·097 | 0·553 | -0·813, 1·1076 |
| **Parent/Guardian of Child Aged** (ref: 4 years and under) | | | |
| 5 to 11 years | -0·335 | 0·327 | -0·881, 0·213 |
| 12 to 16 years | -0·059 | 0·403 | -0·709, 0·627 |
| 17 to 18 years | -0·057 | 0·577 | -0·953, 0·880 |
| Over 18 years | -0·080 | 0·468 | -0·850, 0·693 |
| Not parent/ guardian | | | |
| **Number of Children in Household** (ref: 0) | | | |
| 1 | -0·100 | 0·404 | -0·791, 0·556 |
| 2 | 0·733 | 0·449 | 0·003, 1·478 |
| $\geq$ 3 | 0·328 | 0·465 | -0·432, 1·100 |
| Refused | -0·350 | 0·532 | -1·1235, 0·570 |

Abbreviations: CI = Credible Interval, OR = Odds Ratio, SD = Standard Deviation

Table 11: Posterior Log ORs by covariate for the question “Have you vaccinated your oldest child with one or more doses of the MMR vaccine?”.

| Characteristic | OR | 95% CI |
|---|---|---|
| **Age** (ref: 18–24) | | |
| 25–34 | 0·77 | 0·38, 1·54 |
| 35–44 | 1·26 | 0·61, 2·58 |
| 45–54 | 1·40 | 0·67, 2·92 |
| $\geq$ 55 | 1·72 | 0·76, 3·81 |
| **Gender** (ref: Female) | | |
| Male | 0·63 | 0·43, 0·94 |
| **Social Grade** (ref: High) | | |
| Low | 0·95 | 0·63, 1·42 |
| **Ethnicity** (ref: White) | | |
| Asian | 1·77 | 0·97, 3·51 |
| Black | 0·74 | 0·44, 1·24 |
| Mixed | 0·95 | 0·49, 1·93 |
| Prefer not to say | 1·24 | 0·36, 4·79 |
| **Marital Status** (ref: Married/ Civil Partnership) | | |
| Living as married | 1·18 | 0·57, 2·72 |
| Never Married | 0·74 | 0·45, 1·26 |
| Separated/ Divorced | 0·77 | 0·42, 1·47 |
| Widowed | 0·75 | 0·34, 1·77 |
| **Housing Tenure** (ref: Own) | | |
| Rent-free | 0·32 | 0·11, 1·00 |
| Rent from a housing association | 0·47 | 0·25, 0·89 |
| Rent from a private landlord | 0·48 | 0·29, 0·79 |
| Rent from my local authority | 0·44 | 0·24, 0·83 |
| Family/ friends but pay rent | 0·66 | 0·25, 2·10 |
| Other | 0·79 | 0·27, 2·51 |
| Missing | 1·04 | 0·47, 2·48 |
| **Working Status** (ref: Working full-time) | | |
| Working part-time | 0·73 | 0·44, 1·21 |
| Full-time student | 0·31 | 0·08, 1·15 |
| Not working/ Other | 1·21 | 0·59, 2·52 |
| Retired | 0·67 | 0·35, 1·23 |
| Unemployed | 1·10 | 0·44, 2·93 |
| **Parent/Guardian of Child Aged** (ref: 4 years and under) | | |
| 5 to 11 years | 0·72 | 0·41, 1·24 |
| 12 to 16 years | 0·94 | 0·49, 1·187 |
| 17 to 18 years | 0·94 | 0·39, 2·41 |
| Over 18 years | 0·92 | 0·43, 2·00 |
| **Number of Children in Household** (ref: 0) | | |
| 1 | 0·90 | 0·45, 1·74 |
| 2 | 2·08 | 1·00, 4·38 |
| $\geq$ 3 | 1·39 | 0·65, 3·00 |
| Refused | 0·70 | 0·29, 1·77 |

Abbreviations: CI = Credible Interval, OR = Odds Ratio

Table 12: Posterior ORs by covariate for answer to question "Have you vaccinated your oldest child with one or more doses of the MMR vaccine?".

| Characteristic | Log OR | SD | 95% CI |
|---|---|---|---|
| (Intercept) | 3·226 | 0·647 | 2·001, 4·520 |
| **Age** (ref: 18–24) | | | |
| 25–34 | 0·622 | 0·456 | -0·266, 1·517 |
| 35–44 | -0·191 | 0·461 | -1·126, 0·712 |
| 45–54 | -0·169 | 0·477 | -1·126, 0·783 |
| $\geq$ 55 | 0·449 | 0·515 | -0·555, 1·468 |
| **Gender** (ref: Female) | | | |
| Male | 0·705 | 0·296 | 0·162, 1·283 |
| **Social Grade** (ref: High) | | | |
| Low | -0·212 | 0·281 | -0·790, 0·332 |
| **Ethnicity** (ref: White) | | | |
| Asian | 0·147 | 0·386 | -0·592, 0·935 |
| Black | 0·068 | 0·390 | -0·699, 0·859 |
| Mixed | -0·113 | 0·406 | -0·886, 0·784 |
| Other | -0·267 | 0·827 | -1·1736, 1·361 |
| Prefer not to say | | | |
| **Marital Status** (ref: Married/ Civil Partnership) | | | |
| Living as married | -0·106 | 0·509 | -1·014, 0·938 |
| Never Married | -0·055 | 0·353 | -0·758, 0·631 |
| Separated/ Divorced | -0·474 | 0·446 | -1·327, 0·458 |
| Widowed | -0·459 | 0·519 | -1·1373, 0·608 |
| Missing | -0·429 | 0·791 | -1·807, 1·166 |
| **Housing Tenure** (ref: Own) | | | |
| Rent-free | -0·375 | 0·551 | -1·1428, 0·827 |
| Rent from a housing association | -1·181 | 0·393 | -1·1961, -0·426 |
| Rent from a private landlord | -0·505 | 0·356 | -1·207, 0·209 |
| Rent from my local authority | -1·428 | 0·397 | -2·232, -0·580 |
| Family/ friends but pay rent | -0·300 | 0·595 | -1·407, 0·919 |
| Other | 0·167 | 0·664 | -1·018, 1·583 |
| Missing | -0·094 | 0·561 | -1·122, 1·156 |
| **Working Status** (ref: Working full-time) | | | |
| Working part-time | 0·916 | 0·463 | 0·055, 1·898 |
| Full-time student | 0·739 | 0·687 | -0·502, 2·235 |
| Not working/ Other | 0·108 | 0·431 | -0·711, 0·965 |
| Retired | -0·661 | 0·427 | -1·547, 0·198 |
| Unemployed | -0·177 | 0·465 | -1·039, 0·765 |
| **Parent/Guardian of Child Aged** (ref: 4 years and under) | | | |
| 5 to 11 years | 0·059 | 0·490 | -0·896, 1·027 |
| 12 to 16 years | 1·268 | 0·634 | 0·097, 2·543 |
| 17 to 18 years | 0·275 | 0·711 | -1·029, 1·779 |
| Over 18 years | 0·437 | 0·506 | -0·584, 1·427 |
| Not parent/ guardian | 0·491 | 0·452 | -0·446, 1·328 |
| **Number of Children in Household** (ref: 0) | | | |
| 1 | 0·086 | 0·461 | -0·758, 1·055 |
| 2 | 0·103 | 0·495 | -0·837, 1·100 |
| $\geq$ 3 | 0·424 | 0·639 | -0·716, 1·758 |
| Refused | -0·681 | 0·484 | -1·1594, 0·412 |

Abbreviations: CI = Credible Interval, OR = Odds Ratio, SD = Standard Deviation

Table 13: Posterior Log ORs by covariate for the statement of vaccination effectiveness.

| Characteristic | OR | 95% CI |
|---|---|---|
| **Age** (ref: 18–24) | | |
| 25–34 | 1·86 | 0·77, 4·56 |
| 35–44 | 0·83 | 0·32, 2·04 |
| 45–54 | 0·84 | 0·32, 2·19 |
| $\geq 55$ | 1·57 | 0·57, 4·34 |
| **Gender** (ref: Female) | | |
| Male | 2·02 | 1·18, 3·61 |
| **Social Grade** (ref: High) | | |
| Low | 0·81 | 0·45, 1·39 |
| **Ethnicity** (ref: White) | | |
| Asian | 1·16 | 0·55, 2·55 |
| Black | 1·07 | 0·50, 2·36 |
| Mixed | 0·89 | 0·41, 2·19 |
| Other | 0·77 | 0·18, 3·90 |
| **Marital Status** (ref: Married/ Civil Partnership) | | |
| Living as married | 0·90 | 0·36, 2·56 |
| Never Married | 0·95 | 0·47, 1·88 |
| Separated/ Divorced | 0·62 | 0·27, 1·58 |
| Widowed | 0·63 | 0·25, 1·84 |
| Missing | 0·65 | 0·16, 3·21 |
| **Housing Tenure** (ref: Own) | | |
| Rent-free | 0·69 | 0·24, 2·29 |
| Rent from a housing association | 0·31 | 0·14, 0·65 |
| Rent from a private landlord | 0·60 | 0·30, 1·23 |
| Rent from my local authority | 0·24 | 0·11, 0·56 |
| Family/ friends but pay rent | 0·74 | 0·24, 2·51 |
| Other | 1·18 | 0·36, 4·87 |
| Missing | 0·91 | 0·33, 3·18 |
| **Working Status** (ref: Working full-time) | | |
| Working part-time | 2·50 | 1·06, 6·67 |
| Full-time student | 2·09 | 0·61, 9·35 |
| Not working/ Other | 1·11 | 0·49, 2·62 |
| Retired | 0·52 | 0·21, 1·22 |
| Unemployed | 0·84 | 0·35, 2·15 |
| **Parent/Guardian of Child Aged** (ref: 4 years and under) | | |
| 5 to 11 years | 1·06 | 0·41, 2·79 |
| 12 to 16 years | 3·55 | 1·10, 12·72 |
| 17 to 18 years | 1·32 | 0·36, 5·92 |
| Over 18 years | 1·55 | 0·56, 4·17 |
| Not parent/ guardian | 1·63 | 0·64, 3·77 |
| **Number of Children in Household** (ref: 0) | | |
| 1 | 1·09 | 0·47, 2·87 |
| 2 | 1·11 | 0·43, 3·00 |
| $\geq 3$ | 1·53 | 0·49, 5·80 |
| Refused | 0·51 | 0·20, 1·51 |

Abbreviations: CI = Credible Interval, OR = Odds Ratio



Table 14: Posterior ORs by covariate for the statement of vaccination effectiveness.

| Characteristic | Log OR | SD | 95% CI |
| --- | --- | --- | --- |
| (Intercept) | -0·110 | 0·367 | -0·710, 0·480 |
| **Age** (ref: 18–24) | | | |
| 25–34 | -0·283 | 0·226 | -0·647, 0·101 |
| 35–44 | -0·931 | 0·246 | -1·329, -0·505 |
| 45–54 | -0·746 | 0·270 | -1·169, -0·307 |
| $\geq$ 55 | -1·011 | 0·272 | -1·471, -0·565 |
| **Gender** (ref: Female) | | | |
| Male | 0·104 | 0·116 | -0·085, 0·294 |
| **Social Grade** (ref: High) | | | |
| Low | 0·672 | 0·132 | 0·460, 0·885 |
| **Ethnicity** (ref: White) | | | |
| Asian | 0·673 | 0·172 | 0·390, 0·949 |
| Black | 1·322 | 0·206 | 0·990, 1·672 |
| Mixed | 0·640 | 0·229 | 0·263, 1·019 |
| Other | 0·979 | 0·534 | 0·079, 1·887 |
| Prefer not to say | 0·761 | 0·591 | -0·187, 1·745 |
| **Marital Status** (ref: Married/ Civil Partnership) | | | |
| Living as married | -0·106 | 0·233 | -0·494, 0·280 |
| Never Married | 0·540 | 0·170 | 0·243, 0·816 |
| Separated/ Divorced | 0·213 | 0·239 | -0·195, 0·608 |
| Widowed | 0·437 | 0·328 | -0·098, 0·977 |
| Missing | 0·052 | 0·454 | -0·677, 0·810 |
| **Housing Tenure** (ref: Own) | | | |
| Rent-free | -0·072 | 0·264 | -0·488, 0·368 |
| Rent from a housing association | 0·449 | 0·248 | 0·046, 0·857 |
| Rent from a private landlord | 0·169 | 0·159 | -0·097, 0·439 |
| Rent from my local authority | 0·461 | 0·258 | 0·034, 0·888 |
| Family/ friends but pay rent | -0·035 | 0·295 | -0·517, 0·447 |
| Other | -0·154 | 0·364 | -0·743, 0·480 |
| Missing | 0·700 | 0·245 | 0·312, 1·106 |
| **Working Status** (ref: Working full-time) | | | |
| Working part-time | 0·399 | 0·183 | 0·112, 0·699 |
| Full-time student | 0·006 | 0·320 | -0·519, 0·541 |
| Not working/ Other | -0·197 | 0·221 | -0·546, 0·168 |
| Retired | -0·134 | 0·223 | -0·494, 0·242 |
| Unemployed | -0·021 | 0·253 | -0·437, 0·389 |
| **Parent/Guardian of Child Aged** (ref: 4 years and under) | | | |
| 5 to 11 years | 0·122 | 0·274 | -0·324, 0·577 |
| 12 to 16 years | 0·121 | 0·302 | -0·389, 0·616 |
| 17 to 18 years | 0·019 | 0·478 | -0·761, 0·815 |
| Over 18 years | -0·107 | 0·310 | -0·615, 0·389 |
| Not parent/ guardian | -0·641 | 0·279 | -1·115, -0·194 |
| **Number of Children in Household** (ref: 0) | | | |
| 1 | 0·364 | 0·239 | -0·013, 0·753 |
| 2 | 0·387 | 0·262 | -0·046, 0·819 |
| $\geq$ 3 | 1·383 | 0·330 | 0·858, 1·1915 |
| Refused | 1·827 | 0·343 | 1·282, 2·385 |

Abbreviations: CI = Credible Interval, OR = Odds Ratio, SD = Standard Deviation

Table 15: Posterior Log ORs by covariate for the statement “I am concerned about serious adverse effects (i.e. negative side effects or unwanted results) of vaccines”.

| Characteristic | OR | 95% CI |
|---|---|---|
| **Age** (ref: 18–24) | | |
| 25–34 | 0·75 | 0·52, 1·11 |
| 35–44 | 0·39 | 0·26, 0·60 |
| 45–54 | 0·47 | 0·31, 0·74 |
| $\geq$ 55 | 0·36 | 0·23, 0·57 |
| **Gender** (ref: Female) | | |
| Male | 1·11 | 0·92, 1·34 |
| **Social Grade** (ref: High) | | |
| Low | 1·96 | 1·58, 2·42 |
| **Ethnicity** (ref: White) | | |
| Asian | 1·96 | 1·48, 2·58 |
| Black | 3·75 | 2·69, 5·32 |
| Mixed | 1·90 | 1·30, 2·77 |
| Other | 2·66 | 1·08, 6·60 |
| Prefer not to say | 2·14 | 0·83, 5·73 |
| **Marital Status** (ref: Married/ Civil Partnership) | | |
| Living as married | 0·90 | 0·61, 1·32 |
| Never Married | 1·72 | 1·28, 2·26 |
| Separated/ Divorced | 1·24 | 0·82, 1·84 |
| Widowed | 1·55 | 0·91, 2·66 |
| Missing | 1·05 | 0·51, 2·25 |
| **Housing Tenure** (ref: Own) | | |
| Rent-free | 0·93 | 0·61, 1·44 |
| Rent from a housing association | 1·57 | 1·05, 2·36 |
| Rent from a private landlord | 1·18 | 0·91, 1·55 |
| Rent from my local authority | 1·59 | 1·03, 2·43 |
| Family/ friends but pay rent | 0·97 | 0·60, 1·56 |
| Other | 0·86 | 0·48, 1·62 |
| Missing | 2·01 | 1·37, 3·02 |
| **Working Status** (ref: Working full-time) | | |
| Working part-time | 1·49 | 1·12, 2·01 |
| Full-time student | 1·01 | 0·60, 1·72 |
| Not working/ Other | 0·82 | 0·58, 1·18 |
| Retired | 0·87 | 0·61, 1·27 |
| Unemployed | 0·98 | 0·65, 1·48 |
| **Parent/Guardian of Child Aged** (ref: 4 years and under) | | |
| 5 to 11 years | 1·13 | 0·72, 1·78 |
| 12 to 16 years | 1·13 | 0·68, 1·85 |
| 17 to 18 years | 1·02 | 0·47, 2·26 |
| Over 18 years | 0·90 | 0·54, 1·48 |
| Not parent/ guardian | 0·53 | 0·33, 0·82 |
| **Number of Children in Household** (ref: 0) | | |
| 1 | 1·44 | 0·99, 2·12 |
| 2 | 1·47 | 0·96, 2·27 |
| $\geq$ 3 | 3·99 | 2·36, 6·79 |
| Refused | 6·22 | 3·60, 10·86 |

Abbreviations: CI = Credible Interval, OR = Odds Ratio

Table 16: Posterior ORs by covariate for the statement "I am concerned about serious adverse effects (i.e. negative side effects or unwanted results) of vaccines".

### 2.2. Figures

To evaluate the goodness-of-fit for the Bayesian logistic regression model, we conducted a Posterior Predictive Check (PPC). This diagnostic compares the observed data against simulated datasets generated from the posterior predictive distribution, $y_{\text{rep}}$, to ensure the model successfully captures the underlying patterns in the data. The test statistic, $T(\cdot)$, employed for this diagnostic was the mean sufficient statistics, which, in the context of a binomial model, represents the aggregate proportion of vaccinated individuals. As illustrated in Figures 1 2, 3, 4 and 5, the observed mean was centrally positioned within the distribution of the simulated means $(T(y_{rep}))$. This alignment indicated that the model accurately captured the central tendency of the empirical data and did not exhibit systematic bias in its estimation of overall prevalence. Because the observed statistic resided well within the high-density region of the posterior predictive distribution, we found no evidence of model misfit regarding the average outcome, suggesting that the model is well-calibrated for this population-level summary statistic.

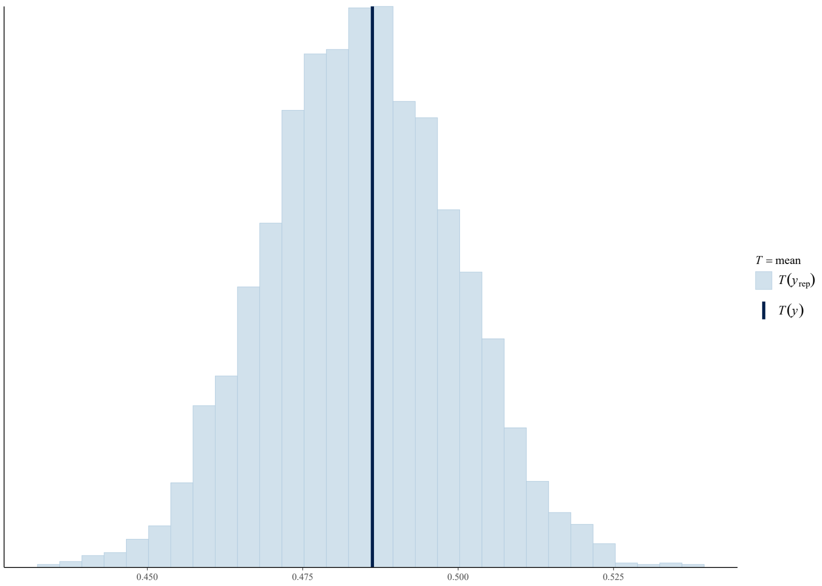


Figure 1: Posterior predictive check for previous year vaccination, comparing the observed mean (dark blue line) to the distribution of means from 4000 posterior predictive simulations (light blue bars).

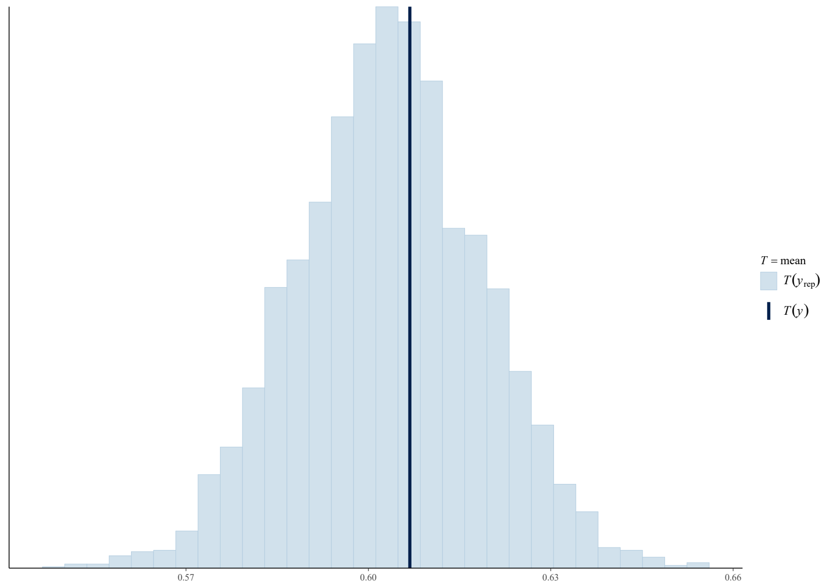


Figure 2: Posterior predictive check for vaccination concern, comparing the observed mean (dark blue line) to the distribution of means from 4000 posterior predictive simulations (light blue bars).

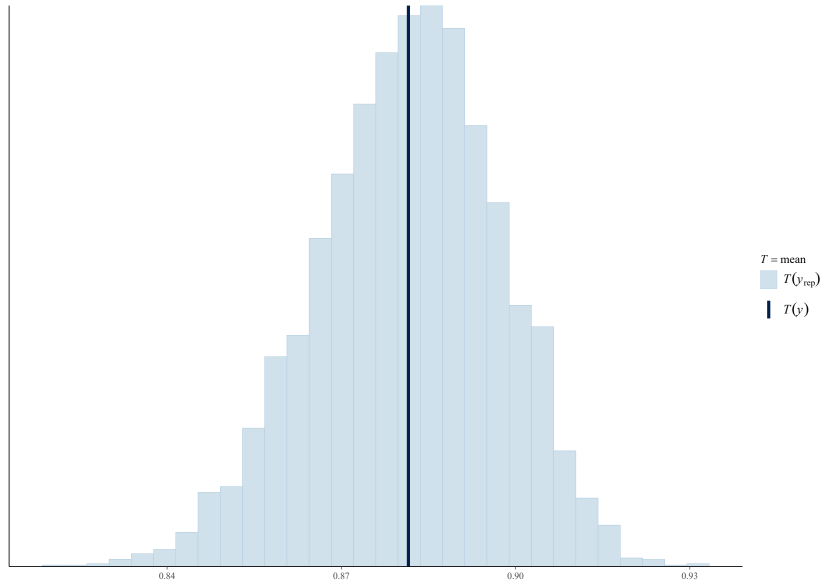


Figure 3: Posterior predictive check for vaccination concern, comparing the observed mean (dark blue line) to the distribution of means from 4000 posterior predictive simulations (light blue bars).

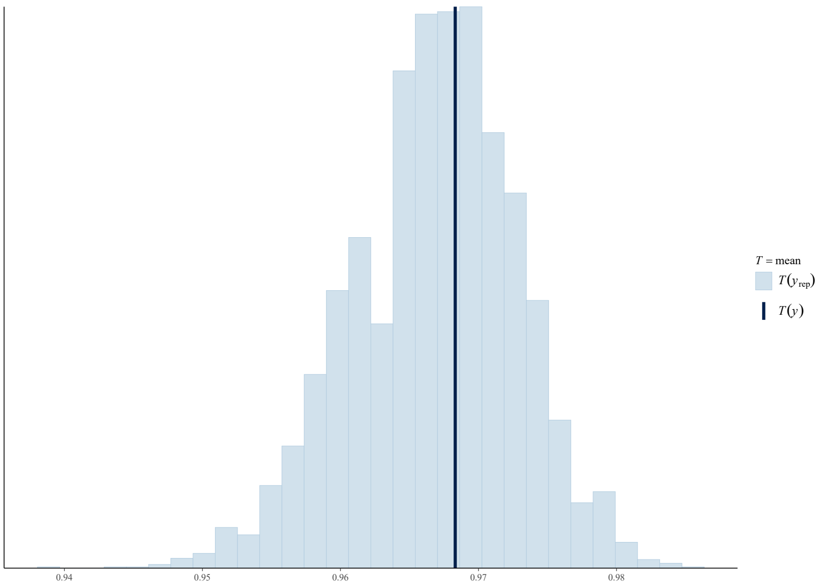


Figure 4: Posterior predictive check for vaccination effectiveness, comparing the observed mean (dark blue line) to the distribution of means from 4000 posterior predictive simulations (light blue bars).

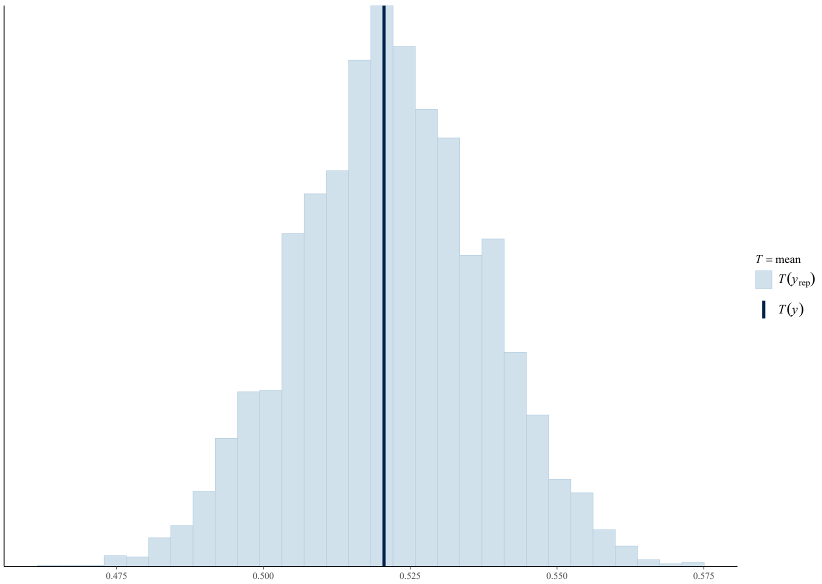


Figure 5: Posterior predictive check for vaccination concern, comparing the observed mean (dark blue line) to the distribution of means from 4000 posterior predictive simulations (light blue bars).
